# Child Developmental Patterns across Subtypes of Hypertensive Disorders of Pregnancy: TMM BirThree Cohort Study

**DOI:** 10.1101/2025.05.02.25326870

**Authors:** Geng Chen, Mami Ishikuro, Hisashi Ohseto, Aoi Noda, Genki Shinoda, Masatsugu Orui, Taku Obara, Shinichi Kuriyama

**Author notes:** Corresponding author Mami Ishikuro Tohoku Medical Megabank Organization, Tohoku University, 2-1 Seiryo-machi, Aoba-ku, Sendai, 980-8573, Japan. The paper (or any part of it) had never been presented or accepted for upcoming presentation at a scientific meeting.

## Abstract

**Background:** Exposure to hypertensive disorders of pregnancy (HDP) and subtypes during the fetal stage has been linked to developmental delays in children. Longitudinal patterns in child development may reveal etiological and pathophysiological mechanisms; however, the associations between HDP subtypes and these patterns remain unclear.

**Objective:** To investigate the association between HDP subtypes and child developmental patterns.

**Study design:** This study included 14,044 mother-child pairs from the Tohoku Medical Megabank Project Birth and Three-Generation Cohort Study. The latent class trajectory model was applied to child development scores at 6, 12, 24, 42, and 48 months of age to generate patterns in five developmental domains: communication, gross motor, fine motor, problem solving, and personal/social. Multinomial Poisson regression analysis adjusted for covariates was used to calculate the risk ratios of different developmental patterns among exposure to HDP and the subtypes among all children and children born at term.

**Results:** Three patterns were identified in respective domains: “normal,” “delay,” “catch-up”. Exposure to HDP was associated with a higher risk of delay pattern in gross motor (risk ratio [RR] 1.29, 95% confidence interval [CI] 1.09-1.52) and of catch-up patterns in fine motor (RR 1.12, 95% CI 1.01-1.24) and problem solving (RR 1.14, 95% CI 1.02-1.28) domains. Exposure to gestational hypertension is associated with a higher risk of catch-up pattern in the fine motor domain (RR 1.17, 95% CI 1.00-1.36). Exposure to preeclampsia (PE) is associated with a higher risk of delay pattern in gross motor, fine motor, and problem solving domains (RR 1.54, 95% CI 1.14–2.04; RR 1.70, 95% CI 1.02–2.65; RR 1.87, 95% CI 1.18–2.81). Children exposed to PE early onset has higher risks of catch-up pattern (RR 1.50, 95% CI 1.01–2.14; RR 1,64, 95% CI 1.05–2.41; RR 1.68, 95% CI 1.15–2.36; RR 1.63, 95% CI 1.11–2.30), and 2–3 folds higher risks of delay pattern (RR 1.93, 95% CI 1.08–3.15; RR 2.12, 95% CI 1.21–3.40; RR 2.70, 95% CI 1.15–5.31; RR 3.00, 95% CI 1.35–5.67) in quadruple domains. In the term-born population, except for PE, there was no association between HDP and adverse patterns.

**Conclusions:** HDP and subtypes, particularly PE early onset, were associated with higher risks of delay child developmental pattern in multiple domains. PE was associated with a higher risk of catch-up pattern. Depending on the HDP subtype, the risks no longer linger in the term-born population.

## 1. Introduction

Hypertensive disorders of pregnancy (HDP) are the most common complications of pregnancy, with a global prevalence of 116.4 per 100000 women of childbearing age.^1^ They are associated with increased risks of mortalities and morbidities in the offspring, while the risks vary among HDP subtypes. Among the phenotypes, preeclampsia (PE), one of the most severe complications of pregnancy, is also associated with increased risk of neurodevelopmental delay, and cardiovascular and metabolic diseases later in life.^2–4^

Child development is a dynamic process spanning from infancy to adulthood. It involves aspects of motor skills, speech and language, social, and performance and cognition.^5^ Early developmental disorders hold the possibility of being persistent toward later years,^6^ while some dysfunctions could dissolve over time.^7^ Although children may exhibit similar performance at certain stages of life, their underlying developmental trajectories may reflect distinct pathological mechanisms rooted in fetal exposures. Identifying patterns in child development could therefore aid in recognizing causative intrauterine factors.

In analyzing repeated measures of child development, prior studies often conducted separate regressions at each time point, thereby overlooking the continuity across time.^8^ Others applied mixed-effect models to examine associations with risk factors, yet these approaches lacked the ability to identify distinct developmental patterns.^9^ In contrast, latent class trajectory models (LCTM) have effectively identified patterns in human development and cognition,^10–16^ supporting their applicability to research on child development and behavior. Moreover, given the distinct pathological processes across HDP subtypes,^17–19^ it is plausible that these differences could induce varied patterns among affected children on their development after birth.

This study aimed to determine the association between HDP and subtypes and developmental patterns in children. We hypothesized that patterns could be identified in child development and that the timing of HDP onset and HDP severity determines the specific pattern.

## 2. Material and methods

### 2.1 Study population

This study was based on the Tohoku Medical Megabank Project Birth and Three-Generation Cohort Study (TMM BirThree Cohort Study). It is a prospective cohort study, recruiting pregnant women from July 2013 to March 2017 in the Miyagi and Iwate Prefectures, Japan. The TMM BirThree Cohort Study continues to collect information on mother-child pairs and their families to contribute to precision medicine.^20–22^ The study protocol was approved by the Ethics Committee of the Tohoku Medical Megabank Organization (2013-1-103-1) on May.27^th^, 2013. All participants provided written informed consent for inclusion in the study.

This study excluded mother–child pairs who met the following criteria: withdrawal of consent (n = 625), abortion (n = 159), stillbirths (n = 185), multiple pregnancies (n = 624), missing HDP subtype (n = 389), and low responders to the Ages and Stages Questionnaires, third edition (ASQ-3) responses (n = 7046) (Figure 1). To build a pattern in the developmental trajectory, we excluded low responders as participants who answered ASQ-3 less than twice. After the exclusions listed above, 14,044 mother–child pairs were included in the analysis.

**Figure 1.**
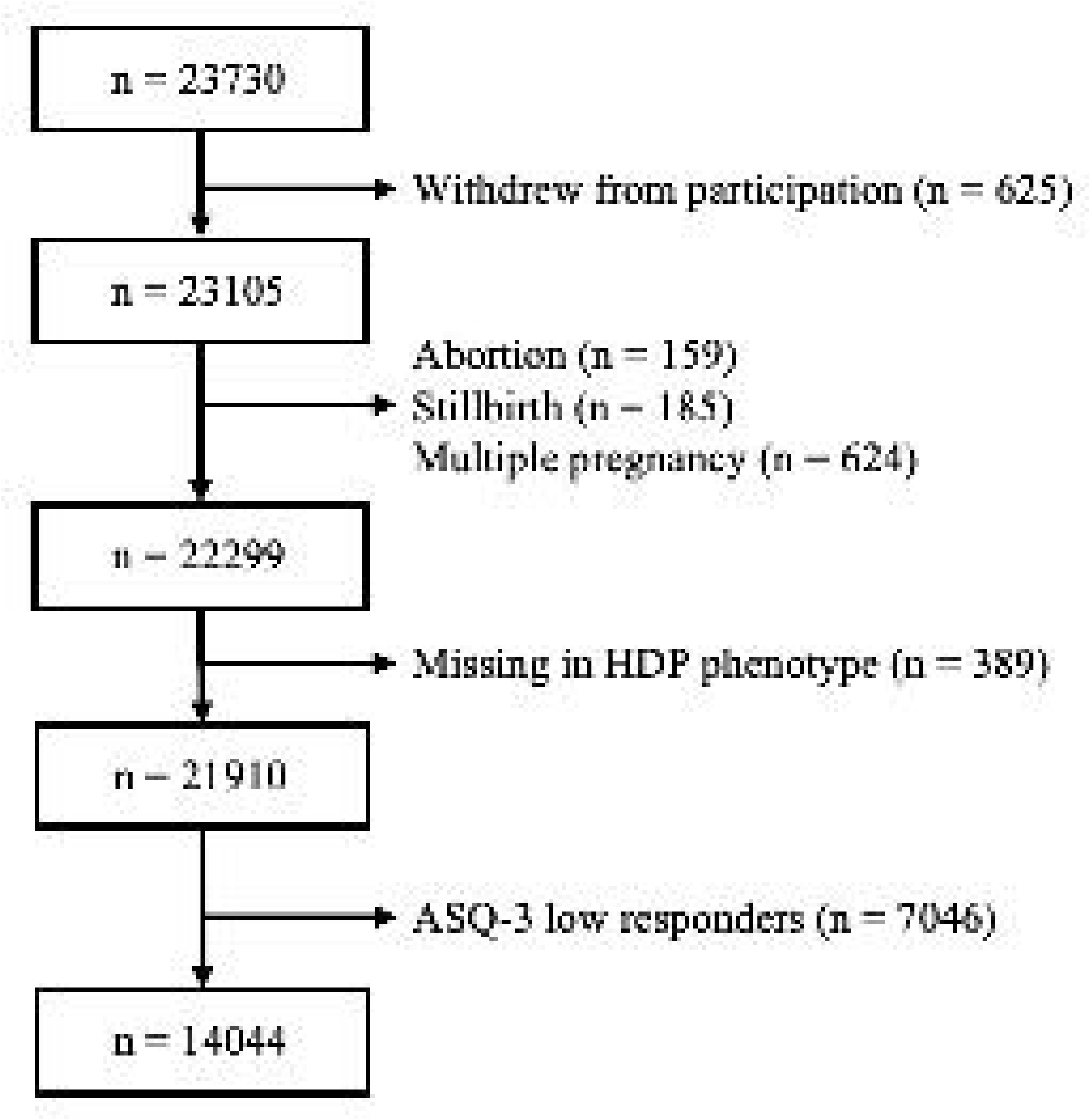
Participant selection flow of the study. HDP, hypertensive disorders of pregnancy; ASQ-3, Age and Stages Questionnaires, third edition.

### 2.2 HDP measures

Participants were identified of HDP subtypes using an algorithm^23^ based on the diagnostic criteria of the American College of Obstetricians and Gynecologists. Gestational hypertension (GH) was defined as systolic blood pressure ≥ 140 mmHg or diastolic blood pressure ≥ 90 mmHg, with no proteinuria during at least one visit after week 20 of gestation in women with previously normal blood pressure. Chronic hypertension (CH) included pregnant women who were hypertensive before 20 weeks of gestation or had a hypertensive disease history before pregnancy. PE was defined as GH with proteinuria (≥ 1+ in the dipstick test) or other PE-related conditions after 20 gestational weeks. Superimposed PE (SPE) was defined as hypertension complicated by proteinuria or other PE-related conditions before 20 weeks of gestation.

Within the PE subtype, participants were further classified based on the timing of proteinuria or other clinical symptoms: PE early onset (PEEO) was defined as onset before 34 weeks of gestation; PE late onset (PELO) was defined as onset at or after 34 weeks of gestation. Subtypes of GH, PE, SPE, and CH were categorized as “HDP-affected,” while not-affected pregnant women were categorized as “HDP-unaffected”.

### 2.3 Child development measures

ASQ-3, a parent-reported handy developmental screening tool targeting five domains: communication, fine motor, gross motor, problem solving, and personal/social,^24^ was used to assess children’s development. Each domain contains six questions about children’s daily activities, with responses of “yes” (10 points), “sometimes” (5 points), or “not yet” (0 points). Responses were summed into a domain score ranging from 0 to 60, with higher scores indicating better performance. The Japanese version of ASQ-3, proven as effective as the original version,^25^ was collected from mothers when their children reached 6, 12, 24, 42, and 48 months of age.

### 2.4 Covariates

Covariates were chosen based on previous studies,^8^ including maternal age at pregnancy (< 35 years old, ≥ 35 years old), family annual income (< 4,000,000 Japanese yen per year, 4,000,000 - < 6,000,000 Japanese yen per year, ≥ 6,000,000 Japanese yen per year), maternal educational attainments (high school or lower, junior or vocational college, university or higher), maternal pre-pregnancy body max index (underweight, < 18.5 kg/m^2^; normal, 18.5 - < 25 kg/m^2^; overweight, ≥ 25 kg/m^2^), parity (primipara, multipara), gestational diabetes mellitus prevalence (yes, no), maternal tobacco use in pregnancy (yes, no), maternal alcohol use in pregnancy (yes, no), maternal folic acid intake in pregnancy (no during pregnancy, yes during pregnancy, yes before and during pregnancy), and child sex (boy, girl). We imputed missing values in the covariates with multiple imputation with chained equations.^26^

### 2.5 Statistically analysis

To identify the developmental patterns in the participant children, ASQ-3 scores at respective time points were z-score scaled to eliminate the artifactual effects caused by different questionnaire contents at every time point and fitted to LCTM.^27^ Linear, quadratic, and cubic models were applied to the fixed effect. Class numbers were given attempts including 2, 3, 4, 5, 6, and 7. We further assessed the model adequacies: the average maximum posterior probability of assignments (APPA) was calculated to ensure a balanced classification, and the odds of correct classification (OCC) were calculated to ensure the overall classification accuracy. The thresholds for good modeling were set as APPA > 70% and OCC > 4.0. Subsequently, the optimal model was selected based on the lowest Bayesian information criterion (BIC). The APPA, OCC, and BIC values for each model are presented in Supplementary Table 1.

Baseline characteristics and developmental patterns were compared between the HDP-affected and HDP-unaffected groups using chi-square tests. We contrasted the rates of different developmental patterns by Poisson regression analysis adjusted for covariates in the following groups: 1) HDP vs. non-HDP, 2) GH vs. non-HDP, 3) PE vs. non-HDP, 4) PEEO vs. non-HDP, and 5) PELO vs. non-HDP. Risk ratios (RR) were calculated along with 95% confidence intervals (CIs). Due to the reported significant association between preterm births and neurodevelopmental disorders,^28^ we conducted the same analysis in a population after excluding preterm births.

### 2.6 Sensitivity analysis

We performed two sensitivity analyses: First, instead of imputing missing values into the covariates, we excluded them and repeated the analysis in a complete case dataset. Second, we excluded children with congenital abnormalities (Supplementary Table 2) at birth or one month of age. Diagnoses were obtained from the medical records of the health checkups.

All analyses were performed using R software (version 4.1.2).^29^ LCTM and model assessments were performed using the R package “lcmm” (version 2.0.2)^30^ and “LCTMtools” (version 0.1.3). Multiple imputation with chained equations was conducted using the R package “mice” (version 3.14.0).^31^ Line graphs of developmental pattern prediction and actual distribution were drawn using the R package “ggplot2” (version 3.3.5).^32^ P value < 0.05 was considered statistically significant.

## 3. Results

### 3.1 Participant characteristics

Table 1 lists the baseline characteristics of the mother-child pairs included in the study. A total of 1,409 mothers (10.0%) experienced HDP during pregnancy, of which 39.2% were affected by GH, 23.3% by CH, 24.8% by PE, and 12.6% by SPE. Within the PE subtype, 85 pregnant women had PEEO, and 265 had PELO. Participants affected by HDP are more likely to be overweight primiparas aged over 35 years, educated below university level, whose family annual income is low, and affected by gestational diabetes mellitus during pregnancy. Their children were more likely to be born preterm. The characteristics among the HDP subtypes are presented in Supplementary Table 3. The results before imputation are shown in Supplementary Tables 4 and 5.

**Table 1.**
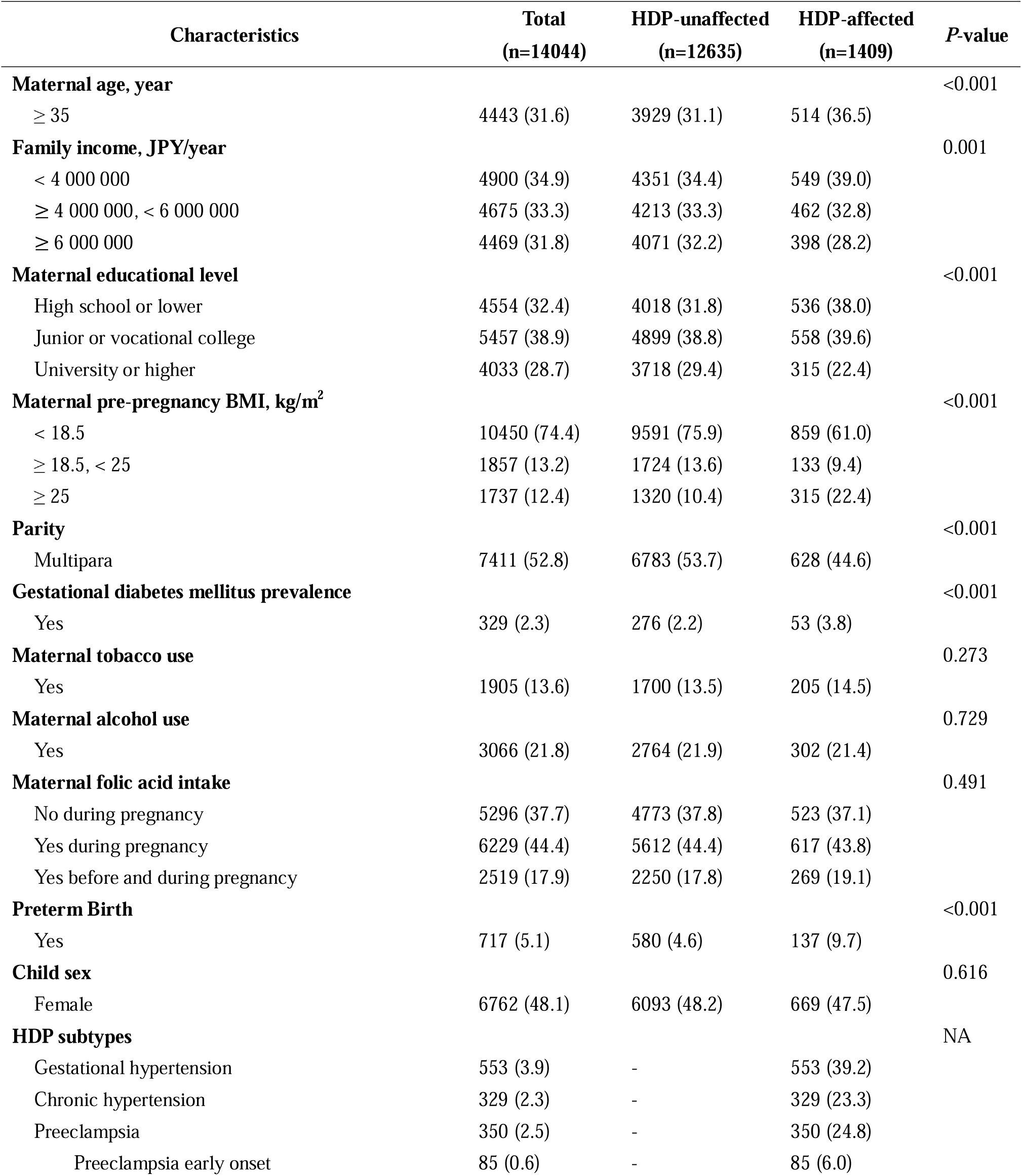
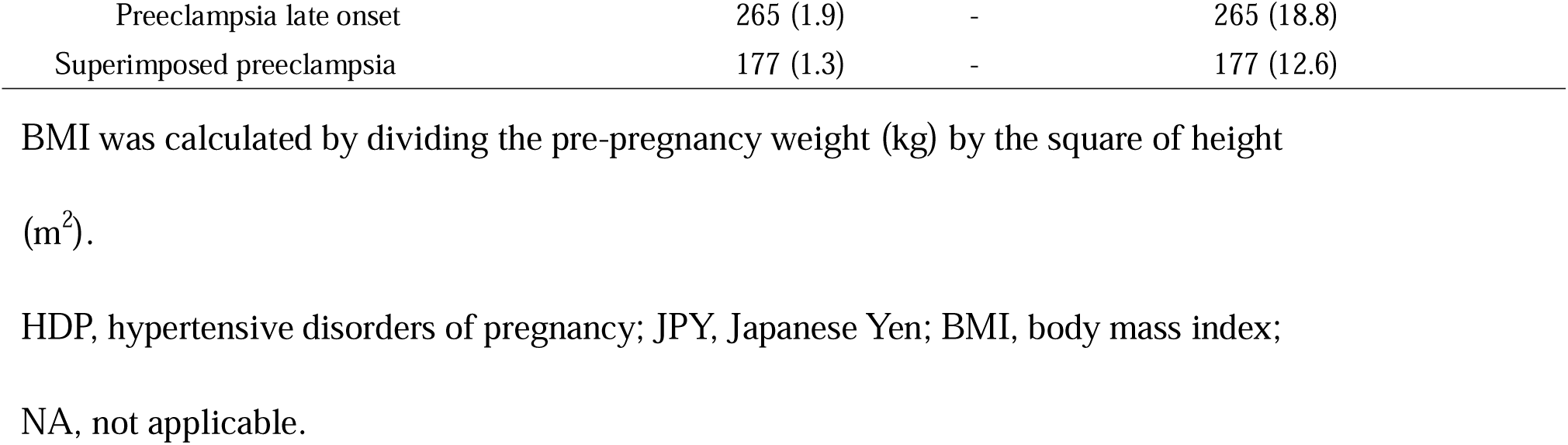
Baseline characteristics of mother–child pairs according to HDP prevalence.

### 3.2 Child developmental patterns

After model adequacies were assessed by APPA and OCC (Supplementary Table 1), the linear model classifying the participant children into three classes by LCTM was optimal, with the lowest BICs in four domains (communication, 145,346.2451; gross motor, 143,839.1751; fine motor, 147,718.4676; problem solving, 142,704.6957; personal/social, 146,730.775). In the fine motor domain, since no model met the OCC threshold, we adopted the same model identified as optimal in the other domains. This model yielded the lowest BIC (147,718.4616), APPAs above the threshold, and acceptable OCC values. Figure 2 shows the prediction graphs of the three developmental patterns in the domains of communication, gross motor, fine motor, problem solving, and personal/social. According to the z-score trends varying with time progression in every pattern, we name the three patterns with “normal,” “catch-up” and “delay”. For every pattern in the five domains, the actual mean and standard deviation values of ASQ-3 scores were in accordance with the prediction graphs (Supplementary Figure 1). Children in the catch-up pattern showed delay signs at first but behave very close to normal at 48 months of age. Children in the delay pattern had persistent delay performances from cuddlers to early childhood.

**Figure 2.**
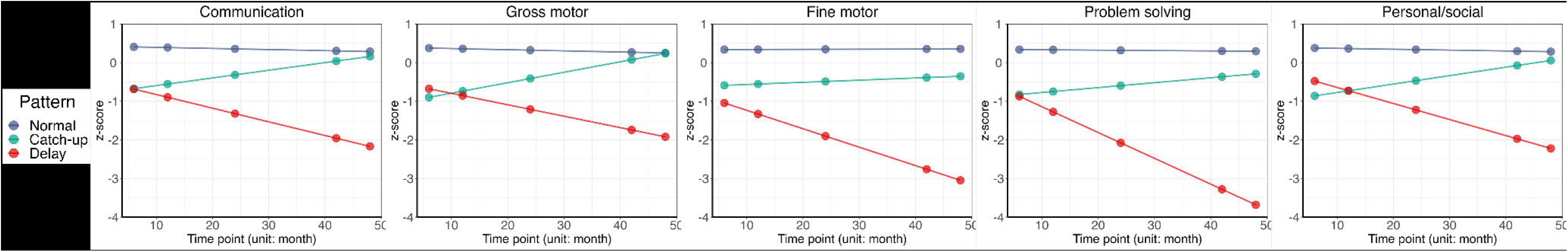
Predicted trajectory of developmental pattern in 5 domains identified by LCTM. Three memberships were given to the participants based on their growing trends in the ASQ-3 z-score: normal (stayed above the mean from 6 to 48 months of age), catch-up (distant from normal at 6 months of age, almost normal at 48 months of age), and delay (distant from normal from 6 to 48 months of age). Each color corresponds to a specific pattern: normal (blue), catch-up (green), or delay (red). From left to right, five developmental domains were presented: communication, gross motor, fine motor, problem solving, and personal/social. LCTM, latent class trajectory model. ASQ-3, Ages and Stages Questionnaires, third edition.

Table 2 presents the distribution of participants for each developmental pattern in the respective domains. Pattern “normal” contained the most participant children, with a percentage ranging from 66.1% to 74.9%. Accordingly, 18.9–26.8% of the participant children were recognized with the pattern “catch-up,” and 3.0–9.1% were recognized with the pattern “delay”. There was a significant difference in the proportion between HDP-affected and HDP-unaffected participants. Children not affected by HDP in fetal life are more likely to be in the pattern “normal” in all five domains.

**Table 2.**
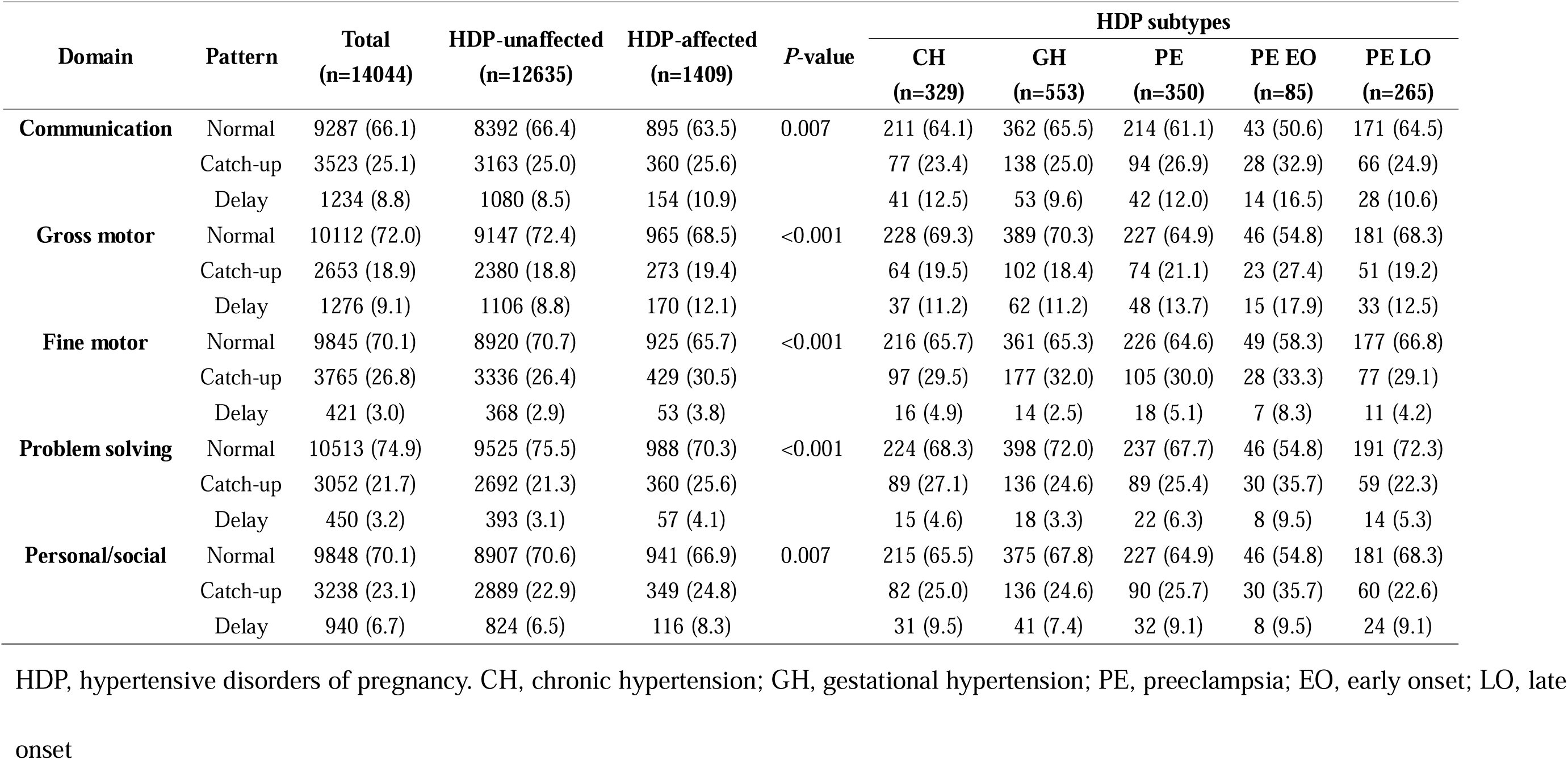
Developmental pattern distribution in HDP subtypes.

| Domain | Pattern | Total<br>(n=14044) | HDP-unaffected<br>(n=12635) | HDP-affected<br>(n=1409) | P-value | HDP subtypes |  |  |  |  |
| --- | --- | --- | --- | --- | --- | --- | --- | --- | --- | --- |
|  |  |  |  |  |  | CH<br>(n=329) | GH<br>(n=553) | PE<br>(n=350) | PE EO<br>(n=85) | PE LO<br>(n=265) |
| **Communication** | Normal | 9287 (66.1) | 8392 (66.4) | 895 (63.5) | 0.007 | 211 (64.1) | 362 (65.5) | 214 (61.1) | 43 (50.6) | 171 (64.5) |
|  | Catch-up | 3523 (25.1) | 3163 (25.0) | 360 (25.6) |  | 77 (23.4) | 138 (25.0) | 94 (26.9) | 28 (32.9) | 66 (24.9) |
|  | Delay | 1234 (8.8) | 1080 (8.5) | 154 (10.9) |  | 41 (12.5) | 53 (9.6) | 42 (12.0) | 14 (16.5) | 28 (10.6) |
| **Gross motor** | Normal | 10112 (72.0) | 9147 (72.4) | 965 (68.5) | <0.001 | 228 (69.3) | 389 (70.3) | 227 (64.9) | 46 (54.8) | 181 (68.3) |
|  | Catch-up | 2653 (18.9) | 2380 (18.8) | 273 (19.4) |  | 64 (19.5) | 102 (18.4) | 74 (21.1) | 23 (27.4) | 51 (19.2) |
|  | Delay | 1276 (9.1) | 1106 (8.8) | 170 (12.1) |  | 37 (11.2) | 62 (11.2) | 48 (13.7) | 15 (17.9) | 33 (12.5) |
| **Fine motor** | Normal | 9845 (70.1) | 8920 (70.7) | 925 (65.7) | <0.001 | 216 (65.7) | 361 (65.3) | 226 (64.6) | 49 (58.3) | 177 (66.8) |
|  | Catch-up | 3765 (26.8) | 3336 (26.4) | 429 (30.5) |  | 97 (29.5) | 177 (32.0) | 105 (30.0) | 28 (33.3) | 77 (29.1) |
|  | Delay | 421 (3.0) | 368 (2.9) | 53 (3.8) |  | 16 (4.9) | 14 (2.5) | 18 (5.1) | 7 (8.3) | 11 (4.2) |
| **Problem solving** | Normal | 10513 (74.9) | 9525 (75.5) | 988 (70.3) | <0.001 | 224 (68.3) | 398 (72.0) | 237 (67.7) | 46 (54.8) | 191 (72.3) |
|  | Catch-up | 3052 (21.7) | 2692 (21.3) | 360 (25.6) |  | 89 (27.1) | 136 (24.6) | 89 (25.4) | 30 (35.7) | 59 (22.3) |
|  | Delay | 450 (3.2) | 393 (3.1) | 57 (4.1) |  | 15 (4.6) | 18 (3.3) | 22 (6.3) | 8 (9.5) | 14 (5.3) |
| **Personal/social** | Normal | 9848 (70.1) | 8907 (70.6) | 941 (66.9) | 0.007 | 215 (65.5) | 375 (67.8) | 227 (64.9) | 46 (54.8) | 181 (68.3) |
|  | Catch-up | 3238 (23.1) | 2889 (22.9) | 349 (24.8) |  | 82 (25.0) | 136 (24.6) | 90 (25.7) | 30 (35.7) | 60 (22.6) |
|  | Delay | 940 (6.7) | 824 (6.5) | 116 (8.3) |  | 31 (9.5) | 41 (7.4) | 32 (9.1) | 8 (9.5) | 24 (9.1) |

HDP, hypertensive disorders of pregnancy. CH, chronic hypertension; GH, gestational hypertension; PE, preeclampsia; EO, early onset; LO, late onset

### 3.3 Risk of outcomes

After adjusting for covariates, compared to HDP-unaffected children, HDP-affected children were associated with a catch-up pattern in fine motor (RR 1.12, 95% CI 1.01–1.24) and problem solving domains (RR 1.14, 95% CI 1.02–1.28), and more associated with a delay pattern in gross motor domain (RR 1.09, 95% CI 1.09–1.52). GH-affected children were associated with a catch-up pattern in the fine-motor domain (RR 1.17, 95% CI 1.00–1.36). PE-affected children were associated with a delay pattern in the gross motor (RR 1.54, 95% CI 1.14–2.04), fine motor (RR 1.70, 95% CI 1.02–2.65), and problem solving domains (RR 1.87, 95% CI 1.18–2.81) (Table 3).

**Table 3.**
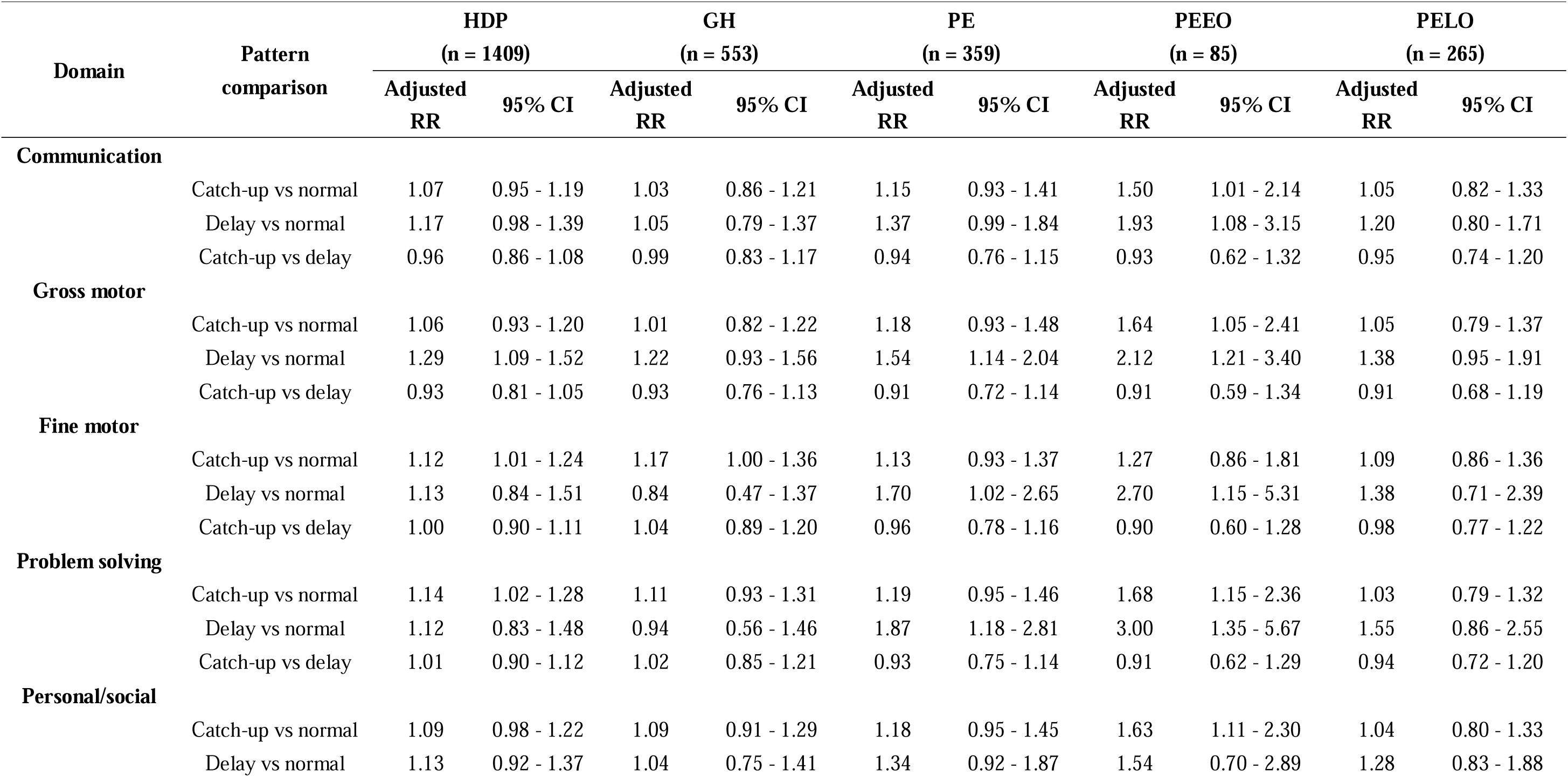
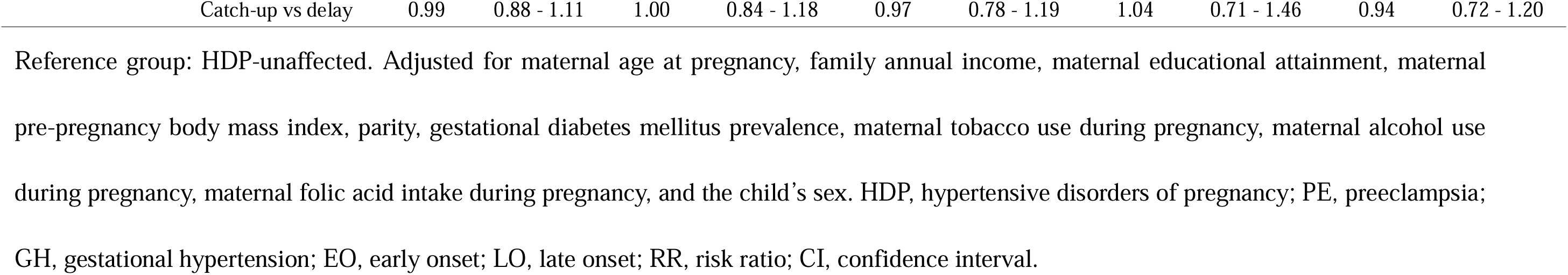
Risk differences of child developmental patterns across HDP and subtypes.

In PE subtypes of onset, compared to HDP-unaffected children, PEEO-affected children had a higher risk of catch-up pattern in communication (RR 1.50, 95% CI 1.01–2.14), gross motor (RR 1.64, 95% CI 1.05–2.41), problem solving (RR 1.68, 95% CI 1.15–2.36) and personal/social domains (RR 1.63, 95% CI 1.11–2.30). Their risks of falling into a delay pattern in communication (RR 1.93, 95% CI 1.08–3.15), gross motor (RR 2.12, 95% CI 1.21–3.40), fine motor (RR 2.70, 95% CI 1.15–5.31), and problem solving domains (RR 3.00, 95% CI 1.35–5.67) were 2–3 folds higher than normal pattern. No significant developmental patterns were observed among the PELO-affected children (Table 3).

When restricted to the term-born population (Table 4), the association is only observed in PE-affected children with a delay pattern in the problem solving domain (RR 1.74, 95% CI 1.02–2.75). There was a similar risk trend for developmental patterns, although some estimates were imprecise. Sensitivity analysis conducted in the complete case population and the congenital abnormality-free population proved the results to be robust (Supplementary Table 6-1, 6-2).

**Table 4.**
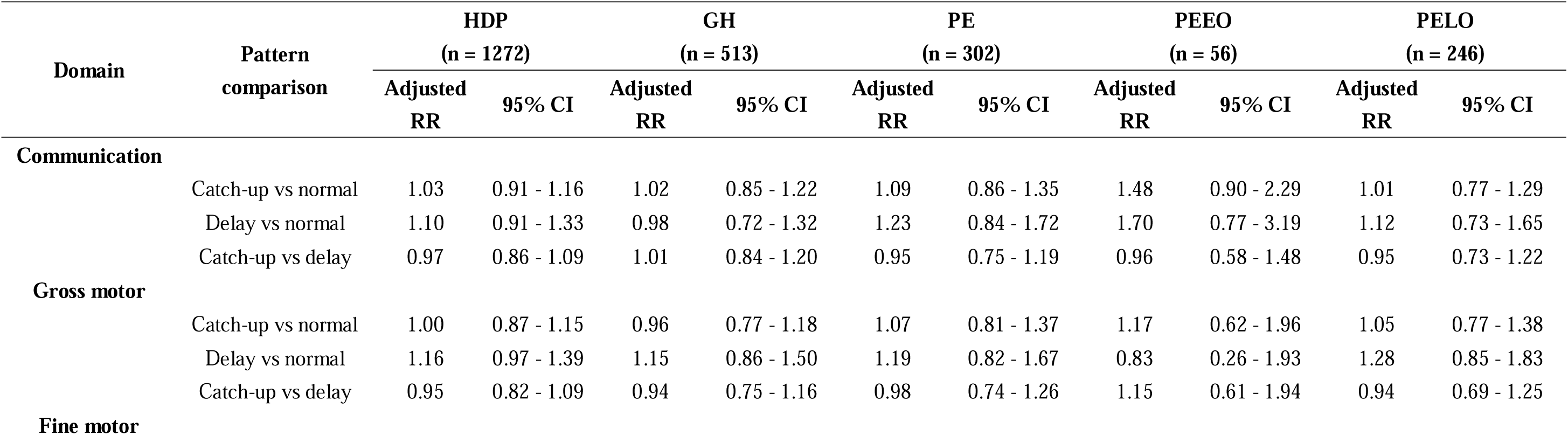
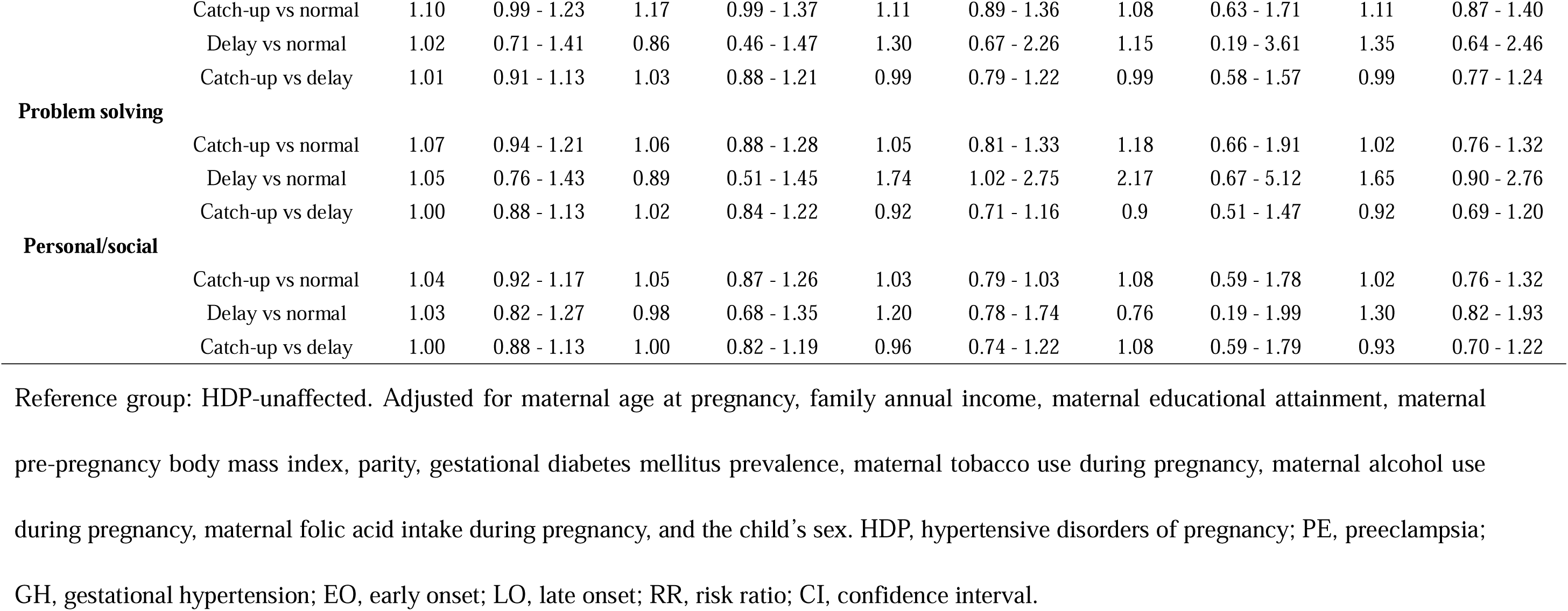
Risk differences of child developmental patterns across HDP and subtypes in term-birth population.

## 4. Comment

### 4.1 Principal Findings

In this study, we identified three child developmental patterns: “normal,” “delay,” and “catch-up” and clarified that exposure to HDP and the subtypes were related to adverse developmental patterns in children. Exposure to the PEEO subtype led to a 2– to 3–fold higher risk of delay pattern in multiple domains. In term-born children, the associations between HDP and adverse developmental patterns were not observed in many domains, except between PE and delay pattern in the problem solving domain.

### 4.2 Results in the Context of What is Known

We found an association between exposure to subtypes of HDP, especially the PE subtype, during the fetal stage and delay child developmental pattern. In mouse models, reduced angiogenesis was reported in the brain cortex of offspring from PE models,^33^ while in humans, reduced fetal brain T2* (tissue oxygenation) and volume have been reported in pregnant women with PE.^34^ Multiple domains are probably affected by PE-induced pathological changes occurring during the fetal period, resulting in an overall impact on fetal brain development.

Previous studies have investigated the risk factors or outcomes of child developmental patterns, identified using the LCTM or its analogous models. Using ASQ-3 questionnaires collected from 3 to 36 months of age in 1,030 children, Li et al.^10^ defined two groups of neuropsychological pattern: “persistently low” and “persistently high,” with group-based trajectory model, although wrongly applied the largest BIC instead of the smallest, and found polyfluoroalkyl substances exposure associated. Black et al. identified four child development patterns: “no problems,” “late socio-emotional problems,” “early cognitive and socioemotional problems,” and “persistent cognitive and socioemotional problems,” using group-based multi trajectory modeling, based on Strength and Difficulty Questionnaire and Bayley Ability Scales II answers from 11,564 children, and found unresolved adverse development impact weight and mental health in adolescence negatively.^11^ McCormick et al. identified five patterns of cognitive development: “high,” “increasing,” “early,” “late,” and “low,” using latent class mixed model on Bayley Scales of Infant and Toddler Development and Wechsler Preschool and Primary Scale of Intelligence from 835 children, and found variables best discriminating the patterns.^35^ In addition to them, we identified new patterns in five respective domains by adhering to ASQ-3 across multiple time points in a large sample size, which ensured consistency in assessment and reliability. To the best of our knowledge, our study is the first to elucidate the association between pregnancy complications and developmental patterns.

### 4.3 Clinical Implications and Research Implications

PEEO, but not PELO, was significantly associated with the delay pattern. Both PEEO and PELO result from placental syncytiotrophoblast stress; however, PEEO is more associated with spiral artery defects, while PELO might be secondary to intervillous malperfusions due to maternal cardiovascular system inabilities.^36^ On the genetic level, dysregulated genes in the placenta are functionally distinct in PEEO and PELO subtypes.^37^ These findings supported the idea that PEEO and PELO affect children’s outcomes through different pathways.

Among the HDP subtypes, we observed an association between PEEO, GH, and the catch-up pattern in child development in multiple domains but not in the term-born population. This may be attributable to the effects of premature birth in small-aged children. After restricting the population to term births, the significant risk of falling into an adverse pattern disappeared in most domains. van Wassenaer et al. followed up on children born after expectant management in cases of fetal growth restriction and PE and found no abnormal behavioral problems.^38^ This indicates that delaying delivery until term helps the offspring maintain a normal developmental pattern. Our findings support the American College of Obstetricians and Gynecologists practice bulletins^39^ that observation until term delivery is recommended in women with preeclampsia without severe features. In terms of population, PE-affected children still had a higher risk of being delay in problem solving. Similar risks have been reported for autism, attention-deficit/hyperactivity disorder, epilepsy, and intellectual disability in children who were exposed to PE but born at term.^40^ Notably, children born to PE-affected mothers exhibit increased resting-state functional connectivity between the amygdala and frontal poles, brain regions implicated in goal-directed behavior. ^41,42^ Thus, their progression in problem solving warrants close monitoring, despite being born at term.

### 4.4 Strengths and Limitations

This study has several strengths. First, it was based on a large-scale cohort study of more than 10,000 mother-child pairs, which ensures low risk of overfitting in the developmental patterns we generated, meanwhile high statistical power in the results. Second, we used LCTM to degrade ASQ-3 data. The LCTM identifies patterns without a detailed definition of cut off values or time points. This enables the inclusion of participants with acceptable missing numbers of ASQ-3 answers, avoiding losses in the exclusion step for ASQ-3 missing answers. Third, the effects of preterm birth on development were ruled out. Fourth, a series of sensitivity analyses were conducted to ensure the robustness of the results.

However, this study has some limitations. In our child developmental pattern, child behaviors after birth, such as education and nutrition, were not considered. Development measurements were included until 48 months of age, which only represented the situations in early childhood. The TMM BirThree Cohort Study will continue to collect developmental data, hoping to generalize developmental patterns that extend to adolescence and adulthood. The TMM BirThree Cohort Study is a regional cohort study conducted in Japan. Therefore, our results should be validated in other regions and ethnic groups. Nevertheless, we inspected developmental patterns under the premise that developmental domains are independent, although evidence indicates that cognitive and motor development are closely associated with each other.^43^ Further studies remain warranted to determine the possible developmental patterns across domains.

### 4.5 Conclusion

HDP and subtypes, particularly PEEO, are associated with a higher risk of delay child developmental pattern. PE was associated with a higher risk of catch-up pattern in multiple domains. Depending on the HDP subtype, the risks no longer linger in the term-born population.

## Glossary

### Author Contributions

Geng Chen: conceptualization, formal analysis, writing – original draft. Mami Ishikuro, Hisashi Ohseto, Shinichi Kuriyama: supervision. Geng Chen, Mami Ishikuro, Hisashi Ohseto, Aoi Noda, Genki Shinoda, Masatsugu Orui, Taku Obara, Shinichi Kuriyama: Data acquisition, interpretation, funding acquisition, writing – review and editing. All authors approved the final submitted manuscript.

## Acknowledgment

The authors would like to thank the Tohoku Medical Megabank Organization, Tohoku University, Iwate Tohoku Medical Megabank Organization, and Iwate Medical University. The full list of members of the Tohoku Medical Megabank Organization is available at https://www.megabank.tohoku.ac.jp/english/a240901/.

## Declaration of generative AI in scientific writing

During the preparation of this work, the authors used ChatGPT 4o to structure some sentences in the writing process. After using this service, the authors reviewed and edited the content as needed and take full responsibility for the content of the published article.

## Data availability

The datasets generated and/or analyzed in the current study are available from the corresponding author upon reasonable request.

## Tweetable statement

Hypertensive disorders of pregnancy and the subtypes were associated with higher risks of adverse patterns in child development. The risks no longer linger in term birth except for preeclampsia.

## Abbreviations

HDP: hypertensive disorders of pregnancy
PE: preeclampsia
PEEO: preeclampsia early onset
PELO: preeclampsia late onset
GH: gestational hypertension
SPE: superimposed preeclampsia
TMM BirThree Cohort Study: the Tohoku Medical Megabank Project Birth and Three-Generation Cohort Study
ASQ-3: Age and Stages Questionnaires third edition
CH: chronic hypertension
APPA: average maximum posterior probability of assignments
OCC: odds of correct classification
BIC: Bayesian information criterion
RR: risk ratio
CI: confidence interval
LCTM: latent class trajectory model

## References

1. Jiang L, Tang K, Magee LA, et al. A global view of hypertensive disorders and diabetes mellitus during pregnancy. Nat Rev Endocrinol. 2022;18(12):760–775. doi: 10.1038/s41574-022-00734-y.

2. Huang C, Wei K, Lee PMY, Qin G, Yu Y, Li J. Maternal hypertensive disorder of pregnancy and mortality in offspring from birth to young adulthood: national population based cohort study. BMJ. 2022;379:e072157. doi: 10.1136/bmj-2022-072157.

3. A. Malhamé I, Nerenberg K, McLaughlin K, Grandi SM, Daskalopoulou SS, Metcalfe A. Hypertensive Disorders and Cardiovascular Severe Maternal Morbidity in the US, 2015-2019. JAMA Netw Open. 2024;7(10):e2436478. doi: 10.1001/jamanetworkopen.2024.36478

3. Pittara T, Vyrides A, Lamnisos D, Giannakou K. Pre-eclampsia and long-term health outcomes for mother and infant: An umbrella review. BJOG. 2021;128(9):1421–1430. doi: 10.1111/1471-0528.16683.

4. Bellman M, Byrne O, Sege R. Developmental assessment of children. Br Med J. 2013;346:e8687. doi: 10.1136/bmj.e8687.

5. Harstad E, Hanson E, Brewster SJ, et al. Persistence of autism spectrum disorder from early childhood through school age. JAMA Pediatr. 2023;177(11):1197–1205. doi: 10.1001/jamapediatrics.2023.4003.

6. Hoeksma MR, Kemner C, Verbaten MN, van Engeland H. Processing capacity in children and adolescents with pervasive developmental disorders. J Autism Dev Disord. 2004;34(3):341–354. doi: 10.1023/b:jadd.0000029555.98493.36.

7. Chen G, Ishikuro M, Ohseto H, et al. Hypertensive disorders of pregnancy, neonatal outcomes and offspring developmental delay in Japan: The Tohoku Medical Megabank Project Birth and Three-Generation Cohort Study. Acta Obstet Gynecol Scand. 2024;103(6):1192–1200. doi: 10.1111/aogs.14820.

8. Melough MM, Li M, Hamra G, et al. Greater Gestational Vitamin D Status is Associated with Reduced Childhood Behavioral Problems in the Environmental Influences on Child Health Outcomes Program. J Nutr. 2023;153(5):1502–1511. doi:10.1016/j.tjnut.2023.03.005

9. Li Q-Q, Huang J, Cai D, et al. Prenatal exposure to legacy and alternative per- and polyfluoroalkyl substances and neuropsychological development trajectories over the first 3 years of life. Environ Sci Technol. 2023;57(9):3746–3757. doi: 10.1021/acs.est.2c07807.

10. Black M, Adjei NK, Strong M, Barnes A, Jordan H, Taylor-Robinson D. Trajectories of child cognitive and socioemotional development and associations with adolescent health in the UK millennium cohort study. J Pediatr. 2023;263:113611. doi: 10.1016/j.jpeds.2023.113611.

11. Luby JL, Belden A, Harms MP, Tillman R, Barch DM. Preschool is a sensitive period for the influence of maternal support on the trajectory of hippocampal development. Proc Natl Acad Sci U S A. 2016;113(20):5742–5747. doi: 10.1073/pnas.1601443113.

12. Mikami M, Hirota T, Adachi M, et al. Trajectories of emotional and behavioral problems in school-age children with coordination difficulties and their relationships to ASD/ADHD traits. Res Dev Disabil. 2023;133:104394. doi: 10.1016/j.ridd.2022.104394.

13. Nagata A, Masumoto T, Nishigori H, et al. Neurodevelopmental outcomes among offspring exposed to corticosteroid and B2-adrenergic agonists in utero. JAMA Netw Open. 2023;6(10):e2339347. doi: 10.1001/jamanetworkopen.2023.39347.

14. Zhou Y, Zhang L, Wang P, et al. Prenatal organophosphate esters exposure and neurodevelopment trajectory in infancy: Evidence from the Shanghai Maternal-Child Pairs Cohort. Sci Total Environ. 2024;927:172366. doi: 10.1016/j.scitotenv.2024.172366.

15. Zhu Z, Chang S, Cheng Y, et al. Early life cognitive development trajectories and intelligence quotient in middle childhood and early adolescence in rural western China. Sci Rep. 2019;9(1):18315. doi: 10.1038/s41598-019-54755-1.

16. Corrêa RRM, Gilio DB, Cavellani CL, et al. Placental morphometrical and histopathology changes in the different clinical presentations of hypertensive syndromes in pregnancy. Arch Gynecol Obstet. 2008;277(3):201–206. doi: 10.1007/s00404-007-0452-z.

17. Maloney KF, Heller D, Baergen RN. Types of maternal hypertensive disease and their association with pathologic lesions and clinical factors. Fetal Pediatr Pathol. 2012;31(5):319–323. doi: 10.3109/15513815.2012.659391.

18. Staff AC. The two-stage placental model of preeclampsia: An update. J Reprod Immunol. 2019;134–135:1-10. doi: 10.1016/j.jri.2019.07.004.

19. Kuriyama S, Yaegashi N, Nagami F, et al. The Tohoku medical Megabank project: Design and mission. J Epidemiol. 2016;26(9):493–511. doi: 10.2188/jea.JE20150268.

20. Sugawara J, Ishikuro M, Obara T, et al. Maternal baseline characteristics and perinatal outcomes: The Tohoku medical Megabank project birth and three-generation cohort study. J Epidemiol. 2022;32(2):69–79. doi: 10.2188/jea.JE20200338.

21. Ishikuro M, Obara T, Osanai T, et al. Strategic methods for recruiting grandparents: The Tohoku medical Megabank birth and three-generation cohort study. Tohoku J Exp Med. 2018;246(2):97–105. doi: 10.1620/tjem.246.97.

22. Mizuno S, Wagata M, Nagaie S, et al. Development of phenotyping algorithms for hypertensive disorders of pregnancy (HDP) and their application in more than 22,000 pregnant women. Sci Rep. 2024;14(1):6292. doi: 10.1038/s41598-024-55914-9.

23. Squires J, Bricker D. ASQ-3 Ages & Stages Questionnaires/ASQ-3 User’s Guide: A Parent-Completed Child-Monitoring System. Baltimore, MD: Brooks Publishing Company; 2009.

24. Mezawa H, Aoki S, Nakayama SF, et al. Psychometric profile of the Ages and Stages Questionnaires, Japanese translation. Pediatr Int. 2019;61(11):1086–1095. doi: 10.1111/ped.13990.

25. Azur MJ, Stuart EA, Frangakis C, Leaf PJ. Multiple imputation by chained equations: What is it and how does it work? Int J Methods Psychiatr Res. 2011;20(1):40–49. doi: 10.1002/mpr.329.

26. Lennon H, Kelly S, Sperrin M, et al. Framework to construct and interpret latent class trajectory modelling. BMJ Open. 2018;8(7):e020683. doi: 10.1136/bmjopen-2017-020683.

27. Mitha A, Chen R, Razaz N, et al. Neurological development in children born moderately or late preterm: National cohort study. BMJ. 2024;384:e075630. doi: 10.1136/bmj-2023-075630.

28. The R Core Team. R: A Language and Environment for Statistical Computing. Vienna, Austria: R Foundation for Statistical Computing; 2018.

29. Proust-Lima C, Philipps V, Liquet B. Estimation of Extended Mixed Models Using Latent Classes and Latent Processes: The R package lcmm. J Stat Softw. 2017;78(2):1–56. doi: 10.18637/jss.v078.i02.

30. van Buuren S, Groothuis-Oudshoorn K. MICE: Multivariate Imputation by Chained Equations in R. Journal of Statistical Software. 2011;45(3):1–67.

31. Wickham H. ggplot2: Elegant Graphics for Data Analysis. Springer-Verlag New York; 2016.

32. Troncoso F, Sandoval H, Ibañez B, et al. Reduced brain cortex angiogenesis in the offspring of the preeclampsia-like syndrome. Hypertension. 2023;80(12):2559–2571. doi: 10.1161/HYPERTENSIONAHA.123.21756.

33. Hall M, de Marvao A, Schweitzer R, et al. Preeclampsia associated differences in the placenta, fetal brain, and maternal heart can be demonstrated antenatally: An observational cohort study using MRI. Hypertension. 2024;81(4):836–847. doi: 10.1161/HYPERTENSIONAHA.123.22442.

34. McCormick BJJ, Caulfield LE, Richard SA, et al. Early life experiences and trajectories of cognitive development. Pediatrics. 2020;146(3):e20193660. doi: 10.1542/peds.2019-3660.

35. Ortega MA, Fraile-Martínez O, García-Montero C, et al. The pivotal role of the placenta in Normal and pathological pregnancies: A focus on preeclampsia, fetal growth restriction, and maternal chronic venous disease. Cells. 2022;11(3):568. doi: 10.3390/cells11030568.

36. Ren Z, Gao Y, Gao Y, et al. Distinct placental molecular processes associated with early-onset and late-onset preeclampsia. Theranostics. 2021;11(10):5028–5044. doi: 10.7150/thno.56141.

37. van Wassenaer AG, Westera J, van Schie PEM, et al. Outcome at 4.5 years of children born after expectant management of early-onset hypertensive disorders of pregnancy. Am J Obstet Gynecol. 2011;204(6):510.e1-510.e9. doi: 10.1016/j.ajog.2011.02.032.

38. American College of Obstetricians and Gynecologists. Gestational hypertension and preeclampsia: ACOG practice bulletin summary, Number 222. Obstet Gynecol. 2020;135(6):1492–1495. doi: 10.1097/AOG.0000000000003892.

39. Sun BZ, Moster D, Harmon QE, Wilcox AJ. Association of Preeclampsia in Term Births With Neurodevelopmental Disorders in Offspring. JAMA Psychiatry. 2020;77(8):823–829. doi:10.1001/jamapsychiatry.2020.0306

40. Mak LE, Croy BA, Kay V, et al. Resting-state functional connectivity in children born from gestations complicated by preeclampsia: A pilot study cohort. Pregnancy Hypertens. 2018;12:23–28. doi:10.1016/j.preghy.2018.02.004

41. Amodio DM, Frith CD. Meeting of minds: the medial frontal cortex and social cognition. Nat Rev Neurosci. 2006;7(4):268–277. doi:10.1038/nrn1884

42. Davis EE, Pitchford NJ, Limback E. The interrelation between cognitive and motor development in typically developing children aged 4-11 years is underpinned by visual processing and fine manual control. Br J Psychol. 2011;102(3):569–584. doi: 10.1111/j.2044-8295.2011.02018.x.

