## supplementary materials for "Child Developmental Patterns across Subtypes of Hypertensive Disorders of Pregnancy: TMM BirThree Cohort Study"

E-mail

***Corresponding author**

Mami Ishikuro

Tohoku Medical Megabank Organization, Tohoku University, 2-1 Seiryo-machi, Aoba-ku, Sendai, 980-8573, Japan.

**Supplementary Figure 1. ASQ-3 score distribution in three developmental patterns in five domains.**

The mean and SDs of the participants’ ASQ-3 scores were calculated by time point and domain and grouped by developmental patterns identified using LCTM. Dots represent the mean values at a given site, whereas vertical bars indicate standard deviations. Each color corresponds to a specific pattern: normal (blue), catch-up (green), or delay (red). From left to right, five developmental domains were presented: communication, gross motor, fine motor, problem solving, and personal/social. The trends in mean values were consistent with the prediction graph (Figure 2). ASQ-3, Ages and Stages Questionnaires, third edition; SD, standard deviation; LCTM, latent class trajectory model.

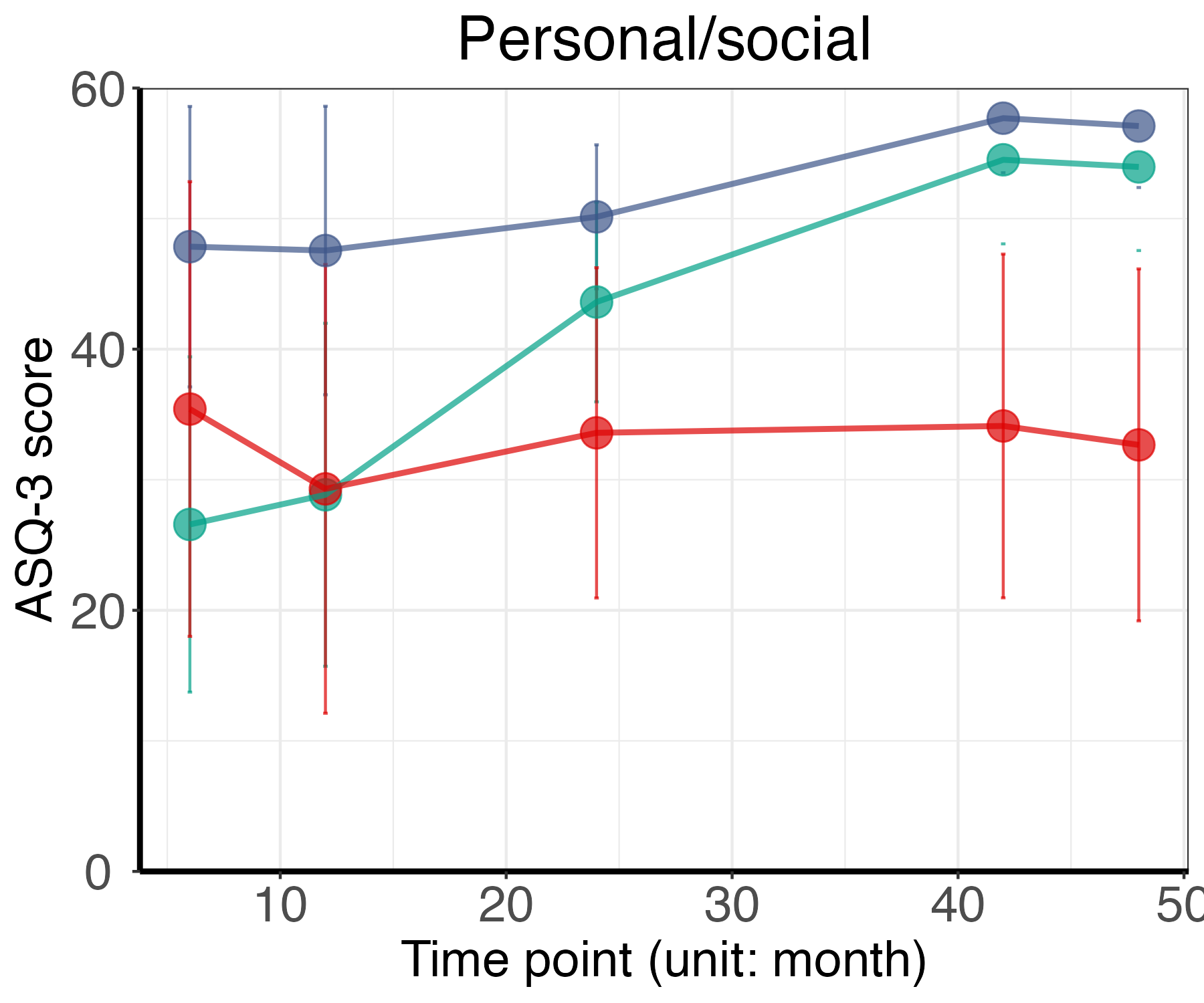

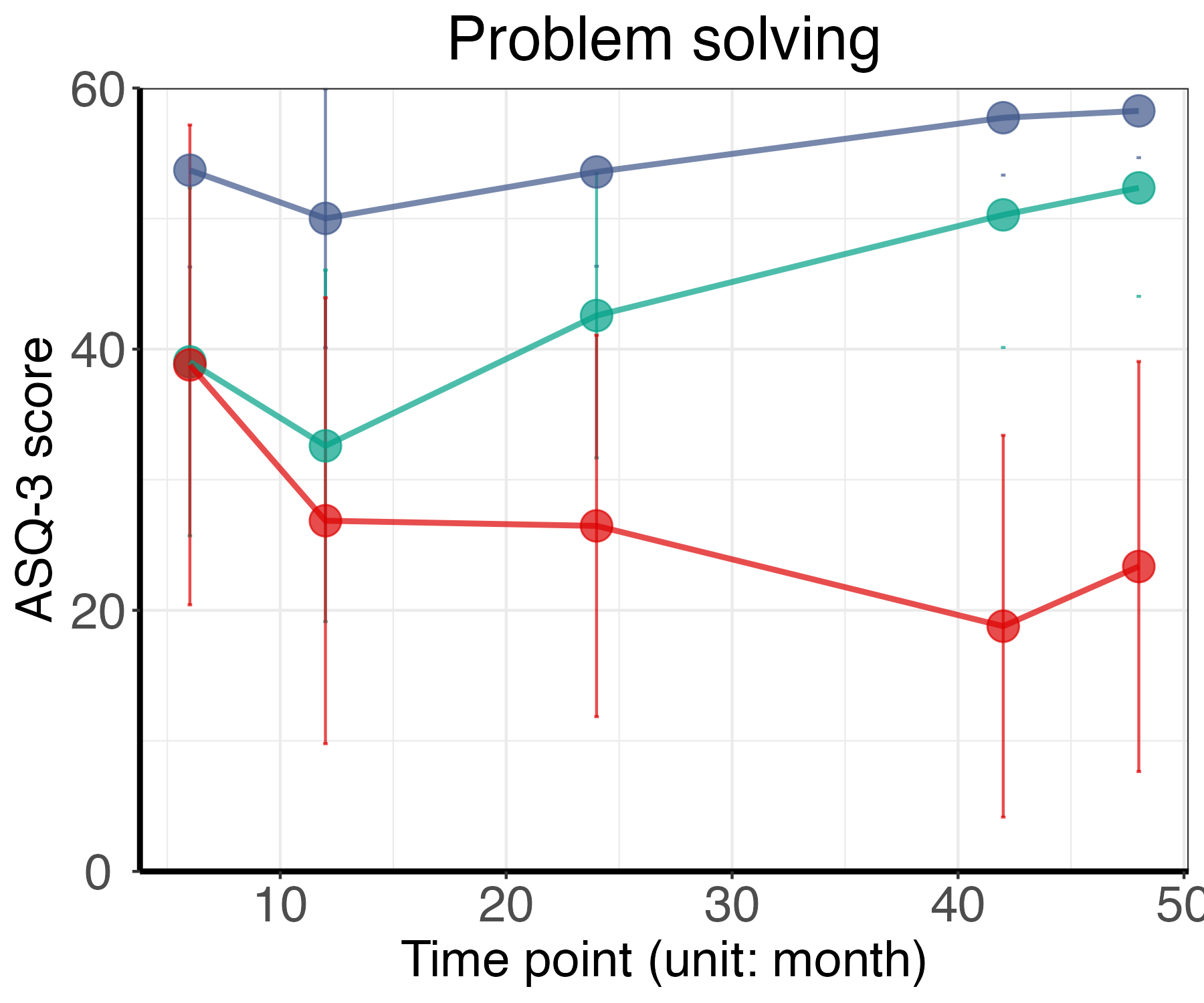

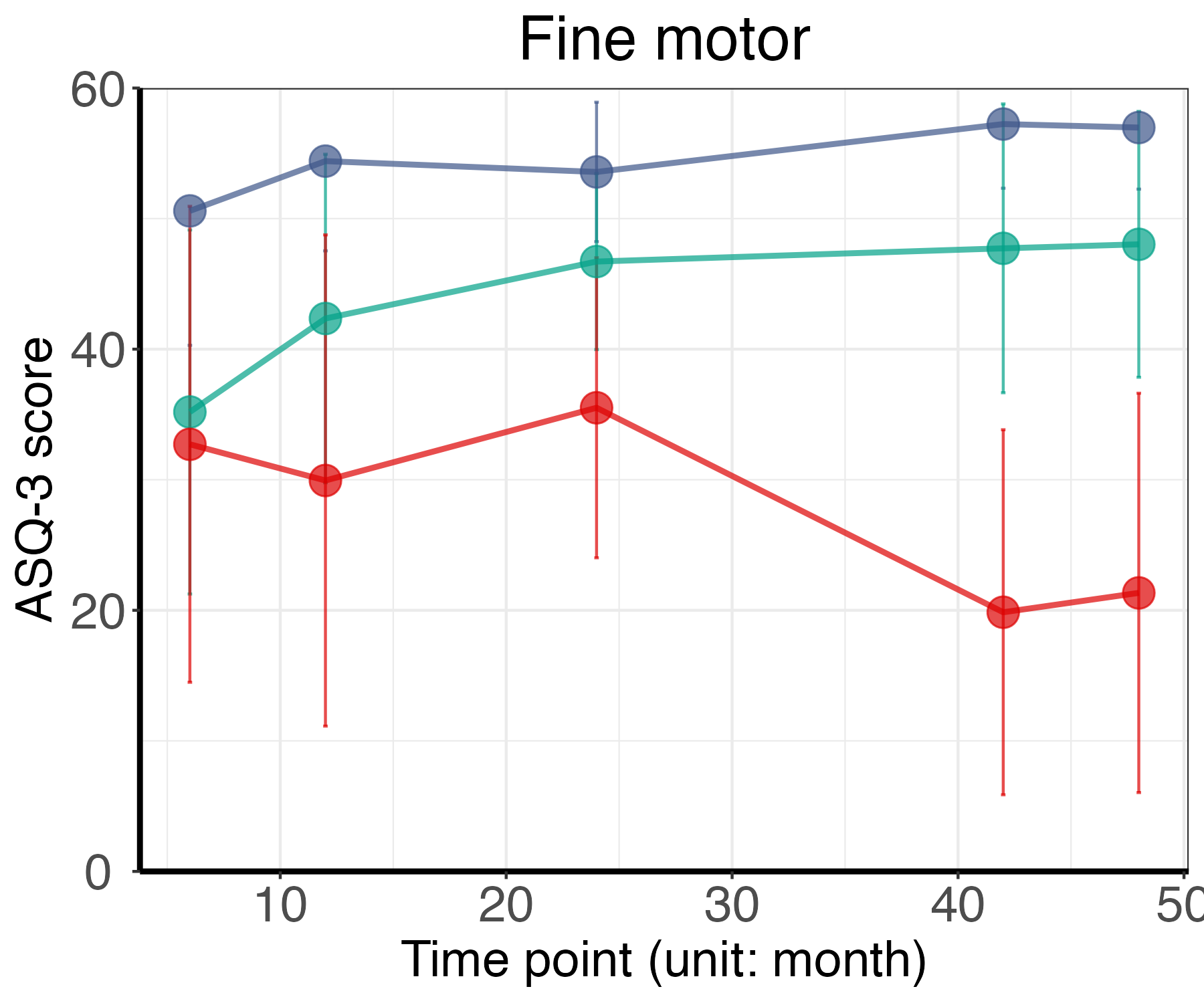

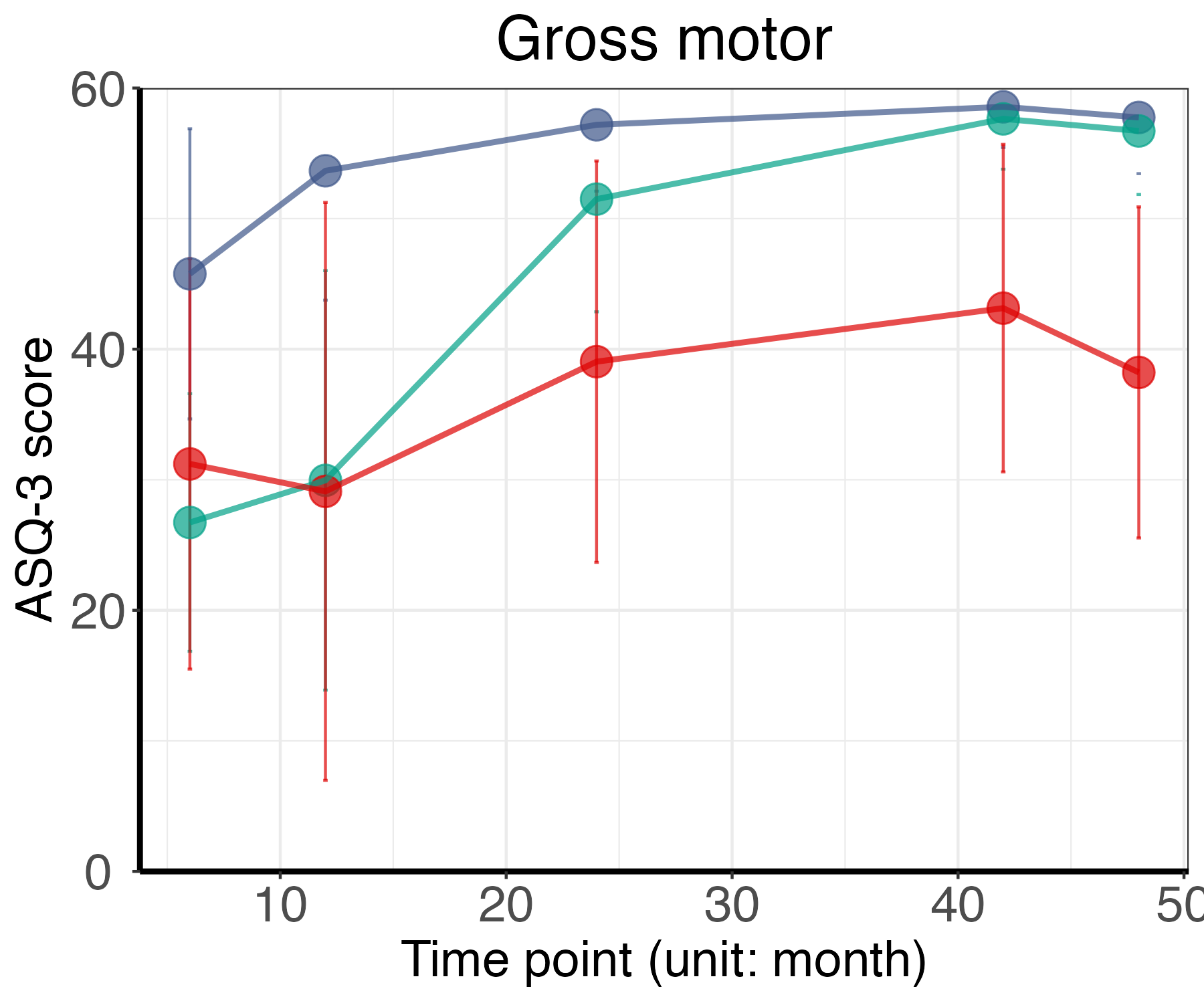

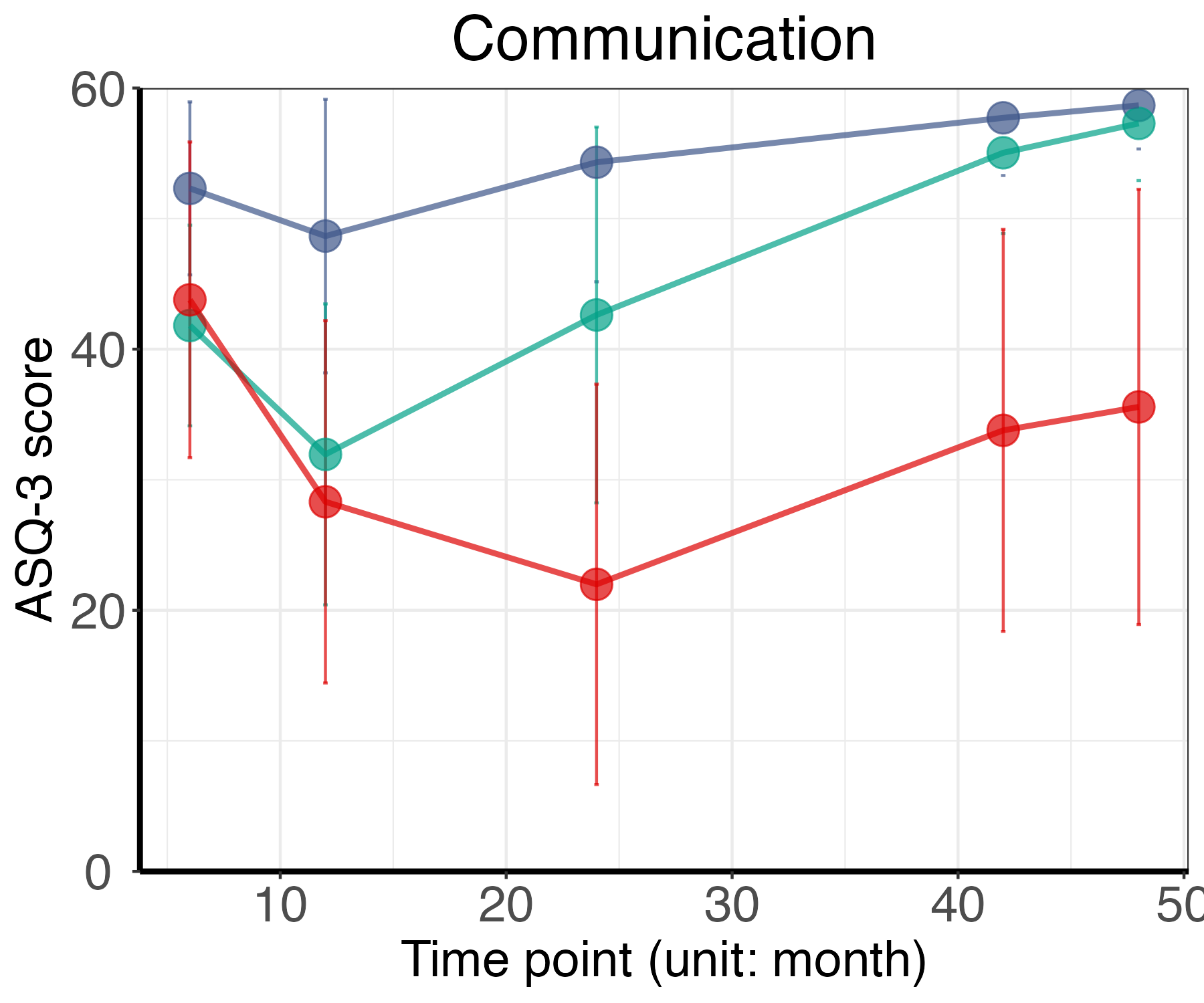

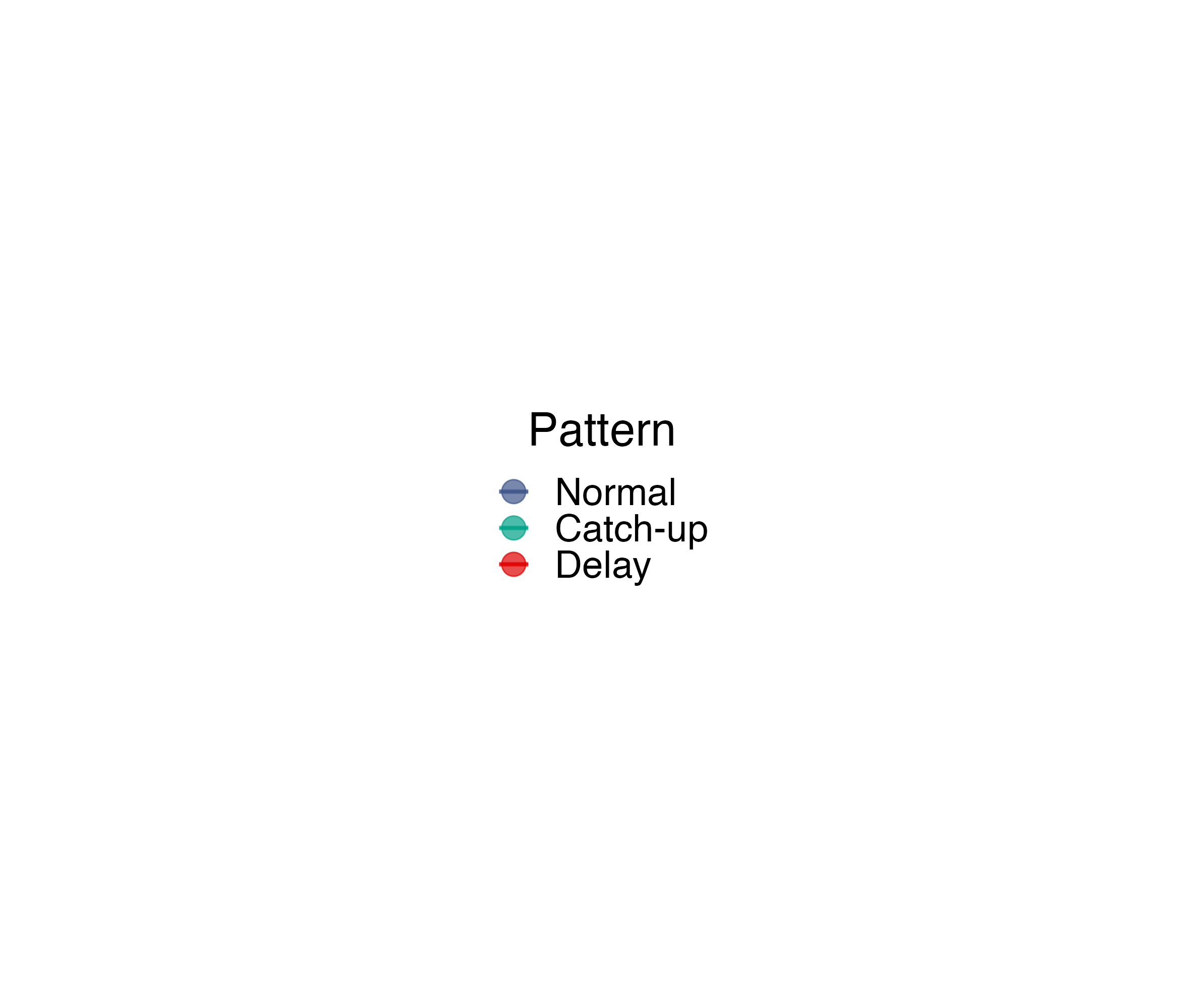

**Supplementary Table 1. Model adequacy assessments in the latent class trajectory model**

| Domain | Class number | Fixed effect | BIC | APPA for every class | | | | | | | OCC for every class | | | | | | |
| --- | --- | --- | --- | --- | --- | --- | --- | --- | --- | --- | --- | --- | --- | --- | --- | --- | --- |
|  |  |  |  | 1 | 2 | 3 | 4 | 5 | 6 | 7 | 1 | 2 | 3 | 4 | 5 | 6 | 7 |
| communication | 2 | linear | 147618.5723 | 0.917 | 0.987 |  |  |  |  |  | 178.695 | 4.737 |  |  |  |  |  |
| communication | 3 | linear | 145346.2451 | 0.867 | 0.877 | 0.729 |  |  |  |  | 52.243 | 4.243 | 7.566 |  |  |  |  |
| communication | 4 | linear | 145077.5274 | 0.876 | 0.69 | 0.862 | 0.783 |  |  |  | 239.898 | 6.223 | 4.258 | 28.465 |  |  |  |
| communication | 5 | linear | 145119.0859 | 0.868 | NA | 0.465 | 0.625 | 0.76 |  |  | 213.448 | NA | 1.98 | 4.689 | 23.213 |  |  |
| communication | 6 | linear | 145138.4517 | 0.838 | 0.608 | 0.704 | NA | 0.573 | 0.616 |  | 245.297 | 4.478 | 47.832 | NA | 2.231 | 14.303 |  |
| communication | 7 | linear | 145075.363 | 0.831 | 0.662 | 0.7 | 0.716 | NA | 0.556 | 0.702 | 296.04 | 5.201 | 78.438 | 89.309 | NA | 2.136 | 36.55 |
| communication | 2 | quadratic | 147625.0244 | 0.918 | 0.987 |  |  |  |  |  | 179.81 | 4.711 |  |  |  |  |  |
| communication | 3 | quadratic | 145354.7053 | 0.868 | 0.877 | 0.732 |  |  |  |  | 53.25 | 4.207 | 7.684 |  |  |  |  |
| communication | 4 | quadratic | 145081.6362 | 0.871 | 0.69 | 0.862 | 0.786 |  |  |  | 228.669 | 6.215 | 4.257 | 28.921 |  |  |  |
| communication | 5 | quadratic | 145080.2611 | 0.854 | 0.676 | 0.861 | 0.72 | 0.678 |  |  | 324.181 | 5.871 | 4.278 | 63.917 | 21.458 |  |  |
| communication | 6 | quadratic | 145149.9201 | 0.842 | 0.702 | 0.578 | NA | 0.478 | 0.621 |  | 243.908 | 47.686 | 4.202 | NA | 2.037 | 13.164 |  |
| communication | 7 | quadratic | 145148.3755 | 0.842 | 0.695 | 0.618 | 0.614 | NA | 0.602 | 0.588 | 263.353 | 59.75 | 37.499 | 4.913 | NA | 2.283 | 16.681 |
| communication | 2 | cubic | 147634.4017 | 0.917 | 0.987 |  |  |  |  |  | 178.299 | 4.724 |  |  |  |  |  |
| communication | 3 | cubic | 145461.2346 | 0.87 | 0.889 | 0.748 |  |  |  |  | 71.794 | 4.227 | 8.46 |  |  |  |  |
| communication | 4 | cubic | 145237.1616 | 0.863 | 0.785 | 0.867 | 0.719 |  |  |  | 226.311 | 37.894 | 3.988 | 7.077 |  |  |  |
| communication | 5 | cubic | 145112.4992 | 0.837 | 0.644 | 0.854 | 0.697 | 0.599 |  |  | 245.424 | 5.416 | 4.154 | 45.013 | 14.156 |  |  |
| communication | 6 | cubic | 145294.4186 | 0.848 | 0.461 | 0.429 | 0.602 | 0.719 | 0.524 |  | 217.247 | 4.571 | 1.797 | 2.979 | 37.537 | 7.953 |  |
| communication | 7 | cubic | 145252.6342 | 0.85 | 0.687 | 0.551 | NA | 0.464 | 0.555 | 0.344 | 233.419 | 39.858 | 15.054 | NA | 3.58 | 2.154 | 4.622 |
| gross motor | 2 | linear | 146470.6846 | 0.9 | 0.978 |  |  |  |  |  | 103.673 | 3.833 |  |  |  |  |  |
| gross motor | 3 | linear | 143839.1751 | 0.872 | 0.903 | 0.74 |  |  |  |  | 50.127 | 4.315 | 11.551 |  |  |  |  |
| gross motor | 4 | linear | 143310.7139 | 0.901 | 0.891 | 0.687 | 0.813 |  |  |  | 541.52 | 4.392 | 9.109 | 27.098 |  |  |  |
| gross motor | 5 | linear | 143339.427 | 0.891 | NA | 0.808 | 0.617 | 0.795 |  |  | 426.246 | NA | 2.943 | 8.388 | 25.417 |  |  |
| gross motor | 6 | linear | 143393.0305 | 0.882 | NA | 0.32 | 0.592 | 0.427 | 0.751 |  | 329.952 | NA | 2.646 | 8.016 | 1.881 | 19.879 |  |
| gross motor | 7 | linear | 143428.5352 | 0.884 | NA | 0.38 | 0.585 | NA | 0.293 | 0.749 | 342.9 | NA | 1.82 | 7.561 | NA | 2.627 | 20.432 |
| gross motor | 2 | quadratic | 146479.8287 | 0.9 | 0.978 |  |  |  |  |  | 103.674 | 3.831 |  |  |  |  |  |
| gross motor | 3 | quadratic | 143851.3033 | 0.872 | 0.903 | 0.744 |  |  |  |  | 51.831 | 4.288 | 11.645 |  |  |  |  |
| gross motor | 4 | quadratic | 143319.1802 | 0.898 | 0.891 | 0.687 | 0.813 |  |  |  | 518.912 | 4.393 | 9.104 | 27.165 |  |  |  |
| gross motor | 5 | quadratic | 143397.8608 | 0.899 | NA | 0.61 | 0.467 | 0.764 |  |  | 337.836 | NA | 6.324 | 1.85 | 18.679 |  |  |
| gross motor | 6 | quadratic | 143464.0317 | 0.863 | 0.583 | 0.308 | NA | 0.387 | 0.736 |  | 198.01 | 6.883 | 2.117 | NA | 1.94 | 17.209 |  |
| gross motor | 7 | quadratic | 143458.0709 | 0.791 | 0.657 | 0.53 | NA | NA | 0.323 | 0.515 | 177.256 | 24.783 | 5.118 | NA | NA | 1.714 | 8.743 |
| gross motor | 2 | cubic | 145793.1471 | 0.877 | 0.925 |  |  |  |  |  | 24.657 | 3.571 |  |  |  |  |  |
| gross motor | 3 | cubic | 143940.1249 | 0.865 | 0.902 | 0.78 |  |  |  |  | 69.539 | 4.093 | 12.289 |  |  |  |  |
| gross motor | 4 | cubic | 143341.5401 | 0.886 | 0.686 | 0.891 | 0.807 |  |  |  | 372.731 | 9.112 | 4.397 | 26.634 |  |  |  |
| gross motor | 5 | cubic | 143424.5432 | 0.889 | NA | 0.499 | 0.613 | 0.764 |  |  | 273.26 | NA | 1.906 | 6.407 | 18.63 |  |  |
| gross motor | 6 | cubic | 143417.735 | 0.897 | 0.345 | 0.537 | NA | 0.459 | 0.739 |  | 329.876 | 3.596 | 8.458 | NA | 1.855 | 18.779 |  |
| gross motor | 7 | cubic | 143451.0869 | 0.881 | 0.643 | 0.385 | NA | 0.312 | 0.342 | 0.714 | 334.666 | 11.973 | 3.526 | NA | 1.728 | 1.99 | 21.358 |
| fine motor | 2 | linear | 148776.028 | 0.866 | 0.9 |  |  |  |  |  | 17.09 | 3.414 |  |  |  |  |  |
| fine motor | 3 | linear | 147718.4676 | 0.859 | 0.877 | 0.81 |  |  |  |  | 146.452 | 3.783 | 9.667 |  |  |  |  |
| fine motor | 4 | linear | 147323.6918 | 0.865 | 0.639 | 0.876 | 0.784 |  |  |  | 300.435 | 9.804 | 3.835 | 16.729 |  |  |  |
| fine motor | 5 | linear | 147362.4479 | 0.865 | 0.586 | NA | 0.513 | 0.746 |  |  | 291.574 | 7.868 | NA | 1.816 | 13.598 |  |  |
| fine motor | 6 | linear | 147361.2757 | 0.832 | 0.446 | 0.572 | NA | 0.571 | 0.63 |  | 210.807 | 9.479 | 1.917 | NA | 7.641 | 14.175 |  |
| fine motor | 7 | linear | 147578.1563 | 0.883 | NA | 0.667 | NA | 0.256 | 0.228 | 0.557 | 313.067 | NA | 11.713 | NA | 1.631 | 1.506 | 6.697 |
| fine motor | 2 | quadratic | 148786.9042 | 0.866 | 0.9 |  |  |  |  |  | 17.134 | 3.409 |  |  |  |  |  |
| fine motor | 3 | quadratic | 147727.2842 | 0.859 | 0.877 | 0.81 |  |  |  |  | 146.581 | 3.782 | 9.666 |  |  |  |  |
| fine motor | 4 | quadratic | 147766.7705 | 0.854 | 0.474 | 0.633 | 0.743 |  |  |  | 122.538 | 2.233 | 2.694 | 7.571 |  |  |  |
| fine motor | 5 | quadratic | 147386.6248 | 0.857 | 0.591 | 0.463 | NA | 0.729 |  |  | 211.646 | 7.864 | 1.741 | NA | 13.43 |  |  |
| fine motor | 6 | quadratic | 147470.6853 | 0.821 | 0.599 | 0.327 | NA | NA | 0.622 |  | 94.238 | 8.135 | 1.594 | NA | NA | 10.206 |  |
| fine motor | 7 | quadratic | 147504.8718 | 0.87 | 0.661 | 0.412 | NA | NA | 0.311 | 0.522 | 226.915 | 15.5 | 4.469 | NA | NA | 1.706 | 7.64 |
| fine motor | 2 | cubic | 148938.662 | 0.862 | 0.901 |  |  |  |  |  | 17.32 | 3.274 |  |  |  |  |  |
| fine motor | 3 | cubic | 147736.3069 | 0.856 | 0.877 | 0.809 |  |  |  |  | 141.462 | 3.782 | 9.647 |  |  |  |  |
| fine motor | 4 | cubic | 147346.9967 | 0.865 | 0.643 | 0.877 | 0.778 |  |  |  | 261.878 | 9.901 | 3.852 | 16.833 |  |  |  |
| fine motor | 5 | cubic | 147425.6353 | 0.843 | 0.588 | 0.479 | NA | 0.692 |  |  | 142.692 | 7.985 | 1.762 | NA | 11.749 |  |  |
| fine motor | 6 | cubic | 147438.5332 | 0.805 | 0.5 | 0.534 | 0.453 | NA | 0.445 |  | 143.894 | 10.586 | 6.643 | 1.77 | NA | 6.299 |  |
| fine motor | 7 | cubic | 147383.4398 | 0.878 | 0.666 | 0.438 | NA | NA | 0.378 | 0.671 | 281.41 | 18.127 | 4.796 | NA | NA | 1.735 | 14.724 |
| problem solving | 2 | linear | 144018.0486 | 0.893 | 0.937 |  |  |  |  |  | 27.976 | 4.408 |  |  |  |  |  |
| problem solving | 3 | linear | 142704.6957 | 0.878 | 0.917 | 0.832 |  |  |  |  | 174.682 | 4.4 | 15.189 |  |  |  |  |
| problem solving | 4 | linear | 142742.8958 | 0.875 | NA | 0.509 | 0.771 |  |  |  | 170.781 | NA | 1.63 | 10.305 |  |  |  |
| problem solving | 5 | linear | 142355.6324 | 0.901 | 0.512 | NA | 0.591 | 0.759 |  |  | 385.94 | 1.67 | NA | 9.634 | 19.008 |  |  |
| problem solving | 6 | linear | 142526.2265 | 0.888 | NA | 0.712 | 0.343 | 0.299 | 0.582 |  | 322.771 | NA | 20.463 | 1.564 | 1.278 | 7.852 |  |
| problem solving | 7 | linear | 142361.2432 | 0.892 | 0.696 | 0.439 | NA | NA | 0.347 | 0.708 | 351.803 | 31.477 | 5.636 | NA | NA | 1.571 | 19.593 |
| problem solving | 2 | quadratic | 144029.2141 | 0.893 | 0.937 |  |  |  |  |  | 27.915 | 4.41 |  |  |  |  |  |
| problem solving | 3 | quadratic | 142711.6741 | 0.879 | 0.917 | 0.831 |  |  |  |  | 175.477 | 4.416 | 15.094 |  |  |  |  |
| problem solving | 4 | quadratic | 142750.0282 | 0.877 | 0.444 | 0.492 | 0.761 |  |  |  | 175.8 | 1.436 | 1.743 | 9.759 |  |  |  |
| problem solving | 5 | quadratic | 142369.9235 | 0.894 | 0.596 | NA | 0.607 | 0.766 |  |  | 320.827 | 9.576 | NA | 1.838 | 19.25 |  |  |
| problem solving | 6 | quadratic | 142540.7498 | 0.887 | 0.715 | NA | NA | 0.324 | 0.586 |  | 313.488 | 21.254 | NA | NA | 1.472 | 7.808 |  |
| problem solving | 7 | quadratic | 142522.8198 | 0.869 | 0.674 | 0.387 | NA | 0.325 | NA | 0.511 | 247.48 | 18.831 | 4.339 | NA | 1.659 | NA | 7.806 |
| problem solving | 2 | cubic | 144290.7292 | 0.89 | 0.935 |  |  |  |  |  | 27.395 | 4.25 |  |  |  |  |  |
| problem solving | 3 | cubic | 142720.7438 | 0.88 | 0.917 | 0.831 |  |  |  |  | 176.201 | 4.418 | 15.062 |  |  |  |  |
| problem solving | 4 | cubic | 142360.2281 | 0.893 | 0.619 | 0.901 | 0.786 |  |  |  | 307.543 | 9.634 | 4.297 | 20.877 |  |  |  |
| problem solving | 5 | cubic | 142388.5623 | 0.89 | NA | 0.504 | 0.577 | 0.755 |  |  | 280.399 | NA | 1.697 | 8.617 | 18.292 |  |  |
| problem solving | 6 | cubic | 142677.9604 | 0.86 | 0.668 | 0.287 | NA | 0.386 | 0.538 |  | 196.965 | 14.908 | 1.626 | NA | 1.6 | 7.246 |  |
| problem solving | 7 | cubic | 142482.3132 | 0.868 | 0.673 | 0.421 | NA | NA | 0.327 | 0.486 | 265.952 | 21.596 | 5.574 | NA | NA | 1.588 | 7.537 |
| personal/social | 2 | linear | 148839.3568 | 0.906 | 0.984 |  |  |  |  |  | 152.886 | 3.967 |  |  |  |  |  |
| personal/social | 3 | linear | 146730.775 | 0.86 | 0.892 | 0.763 |  |  |  |  | 58.396 | 4.213 | 10.092 |  |  |  |  |
| personal/social | 4 | linear | 146332.7569 | 0.903 | 0.682 | 0.881 | 0.792 |  |  |  | 528.214 | 8.347 | 4.237 | 22.699 |  |  |  |
| personal/social | 5 | linear | 146371.7378 | 0.908 | 0.638 | NA | 0.533 | 0.768 |  |  | 526.934 | 6.874 | NA | 1.908 | 20.239 |  |  |
| personal/social | 6 | linear | 146363.3169 | 0.895 | 0.439 | 0.598 | 0.507 | NA | 0.736 |  | 449.68 | 3.86 | 13.317 | 1.973 | NA | 19.613 |  |
| personal/social | 7 | linear | 146394.8767 | 0.891 | 0.55 | 0.302 | NA | NA | 0.547 | 0.701 | 466.647 | 11.172 | 3.698 | NA | NA | 1.935 | 22.225 |
| personal/social | 2 | quadratic | 148848.8025 | 0.906 | 0.984 |  |  |  |  |  | 152.924 | 3.964 |  |  |  |  |  |
| personal/social | 3 | quadratic | 146740.0195 | 0.861 | 0.892 | 0.763 |  |  |  |  | 58.561 | 4.214 | 10.085 |  |  |  |  |
| personal/social | 4 | quadratic | 146343.0123 | 0.903 | 0.881 | 0.68 | 0.785 |  |  |  | 493.355 | 4.238 | 8.339 | 21.785 |  |  |  |
| personal/social | 5 | quadratic | 146394.6694 | 0.874 | 0.614 | 0.477 | NA | 0.744 |  |  | 287.089 | 6.405 | 1.846 | NA | 16.886 |  |  |
| personal/social | 6 | quadratic | 146409.485 | 0.879 | 0.593 | NA | 0.348 | 0.462 | 0.719 |  | 309.362 | 8.879 | NA | 3.037 | 1.937 | 17.256 |  |
| personal/social | 7 | quadratic | 146404.1448 | 0.891 | 0.726 | NA | NA | 0.517 | 0.411 | 0.635 | 508.841 | 26.466 | NA | NA | 4.351 | 1.842 | 16.6 |
| personal/social | 2 | cubic | 148478.9021 | 0.86 | 0.915 |  |  |  |  |  | 18.6 | 3.576 |  |  |  |  |  |
| personal/social | 3 | cubic | 146765.7439 | 0.862 | 0.89 | 0.775 |  |  |  |  | 70.439 | 4.19 | 9.862 |  |  |  |  |
| personal/social | 4 | cubic | 146363.745 | 0.879 | 0.879 | 0.668 | 0.775 |  |  |  | 326.509 | 4.237 | 8.129 | 19.895 |  |  |  |
| personal/social | 5 | cubic | 146405.8789 | 0.877 | 0.629 | 0.566 | NA | 0.751 |  |  | 290.603 | 6.913 | 1.987 | NA | 17.617 |  |  |
| personal/social | 6 | cubic | 146444.6817 | 0.869 | 0.588 | 0.322 | 0.413 | NA | 0.708 |  | 250.794 | 7.63 | 2.526 | 1.942 | NA | 15.659 |  |
| personal/social | 7 | cubic | 146427.7465 | 0.89 | 0.566 | 0.27 | 0.324 | 0.585 | NA | 0.71 | 450.045 | 10.628 | 3.178 | 4.138 | 2.006 | NA | 25.474 |
| APPA, average maximum posterior probability of assignments; OCC, odds of correct classification; BIC, Bayesian information criteria; NA, not available. | | | | | | | | | | | | | | | | | |

**Supplementary Table 2. Congenital abnormalities excluded in the sensitivity analysis**

| **Type** | **Congenital abnormality** |
| --- | --- |
| Cranial abnormalities | anencephaly, microcephaly, hydrocephalus, craniotabes, holoprosencephaly, agenesis of corpus callosum |
| Ocular abnormalities | ablepharon, microphthalmia, anophthalmia, cataract |
| Optic abnormalities | hearing impairment, microtia, atresia of auditory canal, cryptotia, low-set ear |
| Oral abnormalities | cleft lip, cleft palate, cleft lip and palate, prosoposchisis, congenital tooth |
| Chromosomal abnormalities | Down syndrome, Trisomy 18, Trisomy 13, Klinefelter syndrome, Turner syndrome, Prader-Willi syndrome, Trisomy 1q |
| Limb abnormalities | polydactyly, zygodactyly, split-hand malformation, split-foot malformation |
| Thoracic abnormalities | congenital diaphragmatic hernia, pulmonary sequestration, congenital cystic adenomatoid malformation, pulmonary hypoplasia, congenital heart disease, arrhythmia |
| Abdominal abnormalities | umbilical hernia, gastroschisis, congenital esophageal atresia, duodenal atresia, atresia of small intestine, atresia of anus/ atresia ani, anorectal anomaly, inguinal hernia |
| Genitourinary abnormalities | congenital hydronephrosis, multicystic dysplastic kidney, renal agenesis, hypospadias, cryptorchid, nonpalpable testis, bladder exstrophy, cloacal exstrophy, clitoromegaly, vaginal atresia, atypical genitalia |
| Epidermal abnormalities | ≥ 6 large melasma (brown, black, red, white), angioma/hemangioma, epidermolysis bullosa, incontinentia pigmenti |
| Musculo-skeletal abnormalities | myelomeningocele, spina bifida, chondrodysplasia, achondroplasia, osteogenesis imperfecta, arthrogryposis multiplex congenita, hypotonia |
| Others | conjoined fetus, amniotic band syndrome |

**Supplementary Table 3. Baseline characteristics of mother–child pairs in the HDP subtypes**

| **Characteristics** | | **CH** | **GH** | **PE** | **PE EO** | **PE LO** |
| --- | --- | --- | --- | --- | --- | --- |
|  |  | **(n=329)** | **(n=553)** | **(n=350)** | **(n=85)** | **(n=265)** |
| **Maternal age, year** | |  |  |  |  |  |
|  | ≥ 35 | 125 (38.0) | 180 (32.5) | 124 (35.4) | 28 (32.9) | 96 (36.2) |
| **Family income, JPY/year** | |  |  |  |  |  |
|  | < 4 000 000 | 120 (36.5) | 217 (39.2) | 147 (42.0) | 46 (54.1) | 101 (38.1) |
|  | ≥ 4 000 000, < 6 000 000 | 124 (37.7) | 179 (32.4) | 103 (29.4) | 20 (23.5) | 83 (31.3) |
|  | ≥ 6 000 000 | 85 (25.8) | 157 (28.4) | 100 (28.6) | 19 (22.4) | 81 (30.6) |
| **Maternal educational level** | |  |  |  |  |  |
|  | High school or lower | 123 (37.4) | 200 (36.2) | 136 (38.9) | 36 (42.4) | 100 (37.7) |
|  | Junior or vocational college | 134 (40.7) | 231 (41.8) | 135 (38.6) | 31 (36.5) | 104 (39.2) |
|  | University or higher | 72 (21.9) | 122 (22.1) | 79 (22.6) | 18 (21.2) | 61 (23.0) |
| **Maternal pre-pregnancy BMI, kg/m^2^** | |  |  |  |  |  |
|  | < 18.5 | 180 (54.7) | 364 (65.8) | 229 (65.4) | 50 (58.8) | 179 (67.5) |
|  | ≥ 18.5, < 25 | 16 (4.9) | 60 (10.8) | 38 (10.9) | 9 (10.6) | 29 (10.9) |
|  | ≥ 25 | 133 (40.4) | 129 (23.3) | 83 (23.7) | 26 (30.6) | 57 (21.5) |
| **Parity** | |  |  |  |  |  |
|  | Multipara | 158 (48.0) | 247 (44.7) | 141 (40.3) | 35 (41.2) | 106 (40) |
| **Gestational diabetes mellitus prevalence** | |  |  |  |  |  |
|  | Yes | 17 (5.2) | 17 (3.1) | 7 (2.0) | 3 (3.5) | 4 (1.5) |
| **Maternal tobacco use** | |  |  |  |  |  |
|  | Yes | 42 (12.8) | 87 (15.7) | 57 (16.3) | 16 (18.8) | 41 (15.5) |
| **Maternal alcohol use** | |  |  |  |  |  |
|  | Yes | 70 (21.3) | 118 (21.3) | 79 (22.6) | 19 (22.4) | 60 (22.6) |
| **Maternal folic acid intake** | |  |  |  |  |  |
|  | No during pregnancy | 126 (38.3) | 199 (36.0) | 123 (35.1) | 37 (43.5) | 86 (32.5) |
|  | Yes during pregnancy | 131 (39.8) | 252 (45.6) | 162 (46.3) | 39 (45.9) | 123 (46.4) |
|  | Yes before and during pregnancy | 72 (21.9) | 102 (18.4) | 65 (18.6) | 9 (10.6) | 56 (21.1) |
| **Preterm birth** | |  |  |  |  |  |
|  | Yes | 26 (7.9) | 40 (7.2) | 48 (13.7) | 29 (34.1) | 19 (7.2) |
| **Child sex** | |  |  |  |  |  |
|  | Female | 144 (43.8) | 257 (46.5) | 180 (51.4) | 42 (49.4) | 138 (52.1) |
| BMI was calculated by dividing the pre-pregnancy weight (kg) by the square of height (m^2^). | | | | | | |
| CH, chronic hypertension; GH, gestational hypertension; PE, preeclampsia; EO, early onset; LO, late onset; | | | | | | |
| JPY, Japanese Yen; BMI, body mass index. | | | | | | |

**Supplementary Table 4. Baseline characteristics of mother–child pairs before imputation according to HDP prevalence.**

| **Characteristics** | | **Total** | **HDP-unaffected** | **HDP-affected** | ***P*-value** |
| --- | --- | --- | --- | --- | --- |
|  |  | **(n=14044)** | **(n=12635)** | **(n=1409)** |  |
| **Maternal age, year** | |  |  |  | <0.001 |
|  | ≥ 35 | 4443 (31.6) | 3929 (31.1) | 514 (36.5) |  |
| **Family income, JPY/year** | |  |  |  | 0.001 |
|  | < 4 000 000 | 4614 (32.9) | 4106 (32.5) | 508 (36.1) |  |
|  | ≥ 4 000 000, < 6 000 000 | 4415 (31.4) | 3982 (31.5) | 433 (30.7) |  |
|  | ≥ 6 000 000 | 4257 (30.3) | 3883 (30.7) | 374 (26.5) |  |
| **Maternal educational level** | |  |  |  | <0.001 |
|  | High school or lower | 4099 (29.2) | 3605 (28.5) | 494 (35.1) |  |
|  | Junior or vocational college | 4911 (35.0) | 4427 (35.0) | 484 (34.4) |  |
|  | University or higher | 3645 (26.0) | 3361 (26.6) | 284 (20.2) |  |
| **Maternal pre-pregnancy BMI, kg/m^2^** | |  |  |  | <0.001 |
|  | < 18.5 | 10293 (73.3) | 9444 (74.7) | 849 (60.3) |  |
|  | ≥ 18.5, < 25 | 1821 (13.0) | 1694 (13.4) | 127 (9.0) |  |
|  | ≥ 25 | 1708 (12.2) | 1294 (10.2) | 414 (29.4) |  |
| **Parity** | |  |  |  | <0.001 |
|  | Multipara | 7397 (52.7) | 6770 (53.6) | 627 (44.5) |  |
| **Gestational diabetes mellitus prevalence** | |  |  |  | <0.001 |
|  | Yes | 329 (2.3) | 276 (2.2) | 53 (3.8) |  |
| **Maternal tobacco use** | |  |  |  | 0.328 |
|  | Yes | 1880 (13.4) | 1678 (13.3) | 202 (14.3) |  |
| **Maternal alcohol use** | |  |  |  | 0.209 |
|  | Yes | 3033 (21.6) | 2738 (21.7) | 295 (20.9) |  |
| **Maternal folic acid intake** | |  |  |  | 0.601 |
|  | No during pregnancy | 5246 (37.4) | 4727 (37.4) | 519 (36.8) |  |
|  | Yes during pregnancy | 6165 (43.9) | 5556 (44.0) | 609 (43.2) |  |
|  | Yes before and during pregnancy | 2494 (17.8) | 2230 (17.6) | 264 (18.7) |  |
| **Preterm birth** | |  |  |  | <0.001 |
|  | Yes | 717 (5.1) | 580 (4.6) | 137 (9.7) |  |
| **Child sex** | |  |  |  | 0.616 |
|  | Female | 6762 (48.1) | 6093 (48.2) | 669 (47.5) |  |
| **HDP subtypes** | |  |  |  | NA |
|  | Gestational hypertension | 553 (3.9) | - | 553 (39.2) |  |
|  | Chronic hypertension | 329 (2.3) | - | 329 (23.3) |  |
|  | Preeclampsia | 350 (2.5) | - | 350 (24.8) |  |
|  | Preeclampsia early onset | 85 (0.6) | - | 85 (6.0) |  |
|  | Preeclampsia late onset | 265 (1.9) | - | 265 (18.8) |  |
|  | Superimposed preeclampsia | 177 (1.3) | - | 177 (12.6) |  |
| BMI was calculated by dividing the pre-pregnancy weight (kg) by the square of height (m^2^). | | | | | |
| HDP, hypertensive disorders of pregnancy; JPY, Japanese Yen; BMI, body mass index; NA, not applicable. | | | | | |

**Supplementary Table 5. Baseline characteristics of the participant mother–child pairs before imputation in the HDP subtypes**

| **Characteristics** | | **CH** | **GH** | **PE** | **PE EO** | **PE LO** |
| --- | --- | --- | --- | --- | --- | --- |
|  |  | **(n=329)** | **(n=553)** | **(n=350)** | **(n=85)** | **(n=265)** |
| **Maternal age, year** | |  |  |  |  |  |
|  | ≥ 35 | 125 (38.0) | 180 (32.5) | 124 (35.4) | 28 (32.9) | 96 (36.2) |
| **Family income, JPY/year** | |  |  |  |  |  |
|  | < 4 000 000 | 110 (33.4) | 198 (35.8) | 137 (39.1) | 41 (48.2) | 96 (36.2) |
|  | ≥ 4 000 000, < 6 000 000 | 118 (35.9) | 164 (29.7) | 98 (28.0) | 19 (22.4) | 79 (29.8) |
|  | ≥ 6 000 000 | 82 (24.9) | 146 (26.4) | 93 (26.6) | 16 (18.8) | 77 (29.1) |
| **Maternal educational level** | |  |  |  |  |  |
|  | High school or lower | 112 (34.0) | 182 (32.9) | 129 (36.9) | 35 (41.2) | 94 (35.5) |
|  | Junior or vocational college | 118 (35.9) | 198 (35.8) | 116 (33.1) | 23 (27.1) | 93 (35.1) |
|  | University or higher | 68 (20.7) | 104 (18.8) | 73 (20.9) | 16 (18.8) | 57 (21.5) |
| **Maternal pre-pregnancy BMI, kg/m^2^** | |  |  |  |  |  |
|  | < 18.5 | 177 (53.8) | 361 (65.3) | 226 (64.6) | 49 (57.6) | 177 (66.8) |
|  | ≥ 18.5, < 25 | 15 (4.6) | 60 (10.8) | 36 (10.3) | 8 (9.4) | 28 (10.6) |
|  | ≥ 25 | 133 (40.4) | 129 (23.3) | 81 (23.1) | 25 (29.4) | 56 (21.1) |
| **Parity** | |  |  |  |  |  |
|  | Multipara | 158 (48.0) | 247 (44.7) | 140 (40.0) | 34 (40.0) | 106 (40.0) |
| **Gestational diabetes mellitus prevalence** | |  |  |  |  |  |
|  | Yes | 17 (5.2) | 17 (3.1) | 7 (2.0) | 3 (3.5) | 4 (1.5) |
| **Maternal tobacco use** | |  |  |  |  |  |
|  | Yes | 41 (12.5) | 86 (15.6) | 56 (16.0) | 16 (18.8) | 40 (15.1) |
| **Maternal alcohol use** | |  |  |  |  |  |
|  | Yes | 70 (21.3) | 115 (20.8) | 76 (21.7) | 19 (22.4) | 57 (21.5) |
| **Maternal folic acid intake** | |  |  |  |  |  |
|  | No during pregnancy | 124 (37.7) | 198 (35.8) | 122 (34.9) | 36 (42.4) | 86 (32.5) |
|  | Yes during pregnancy | 131 (39.8) | 247 (44.7) | 159 (45.4) | 39 (45.9) | 120 (45.3) |
|  | Yes before and during pregnancy | 72 (21.9) | 100 (18.1) | 64 (18.3) | 9 (10.6) | 55 (20.8) |
| **Preterm birth** | |  |  |  |  |  |
|  | Yes | 26 (7.9) | 40 (7.2) | 48 (13.7) | 29 (34.1) | 19 (7.2) |
| **Child sex** | |  |  |  |  |  |
|  | Female | 144 (43.8) | 257 (46.5) | 180 (51.4) | 42 (49.4) | 138 (52.1) |
| BMI was calculated by dividing the pre-pregnancy weight (kg) by the square of height (m^2^). | | | | | | |
| CH, chronic hypertension; GH, gestational hypertension; PE, preeclampsia; EO, early onset; LO, late onset; | | | | | | |
| JPY, Japanese Yen; BMI, body mass index. | | | | | | |

**Supplementary Table 6-1. Risk differences in child developmental patterns across HDP and subtypes in complete cases**

| **Domain** | **Pattern comparison** | **HDP** | | **GH** | | **PE** | | **PEEO** | | **PELO** | |
| --- | --- | --- | --- | --- | --- | --- | --- | --- | --- | --- | --- |
|  |  | **Adjusted RR** | **95% CI** | **Adjusted RR** | **95% CI** | **Adjusted RR** | **95% CI** | **Adjusted RR** | **95% CI** | **Adjusted RR** | **95% CI** |
| **Communication** |  |  |  |  |  |  |  |  |  |  |  |
|  | Catch-up vs normal | 1.07 | 0.95 - 1.21 | 1.07 | 0.89 - 1.28 | 1.14 | 0.90 - 1.41 | 1.54 | 0.98 - 2.29 | 1.03 | 0.78 - 1.33 |
|  | Delay vs normal | 1.14 | 0.93 - 1.37 | 1.03 | 0.74 - 1.39 | 1.41 | 0.99 - 1.94 | 1.88 | 0.94 - 3.32 | 1.29 | 0.85 - 1.86 |
|  | Catch-up vs delay | 0.98 | 0.87 - 1.11 | 1.02 | 0.84 - 1.22 | 0.93 | 0.74 - 1.16 | 0.96 | 0.61 - 1.42 | 0.92 | 0.70 - 1.19 |
| **Gross motor** |  |  |  |  |  |  |  |  |  |  |  |
|  | Catch-up vs normal | 1.07 | 0.93 - 1.23 | 0.99 | 0.79 - 1.23 | 1.16 | 0.89 - 1.49 | 1.43 | 0.84 - 2.26 | 1.09 | 0.80 - 1.45 |
|  | Delay vs normal | 1.27 | 1.05 - 1.52 | 1.28 | 0.96 - 1.67 | 1.49 | 1.06 - 2.02 | 1.78 | 0.89 - 3.15 | 1.41 | 0.95 - 2.00 |
|  | Catch-up vs delay | 0.94 | 0.81 - 1.08 | 0.90 | 0.72 - 1.12 | 0.92 | 0.70 - 1.17 | 0.91 | 0.53 - 1.44 | 0.92 | 0.67 - 1.21 |
| **Fine motor** |  |  |  |  |  |  |  |  |  |  |  |
|  | Catch-up vs normal | 1.09 | 0.97 - 1.22 | 1.17 | 0.99 - 1.38 | 1.11 | 0.89 - 1.37 | 1.18 | 0.73 - 1.78 | 1.09 | 0.85 - 1.38 |
|  | Delay vs normal | 1.08 | 0.77 - 1.49 | 0.80 | 0.41 - 1.40 | 1.67 | 0.95 - 2.72 | 2.42 | 0.86 - 5.28 | 1.45 | 0.72 - 2.58 |
|  | Catch-up vs delay | 1.01 | 0.90 - 1.13 | 1.04 | 0.88 - 1.23 | 0.96 | 0.77 - 1.18 | 0.90 | 0.56 - 1.35 | 0.98 | 0.76 - 1.23 |
| **Problem solving** |  |  |  |  |  |  |  |  |  |  |  |
|  | Catch-up vs normal | 1.11 | 0.98 - 1.26 | 1.12 | 0.92 - 1.35 | 1.20 | 0.95 - 1.50 | 1.66 | 1.06 - 2.44 | 1.07 | 0.81 - 1.40 |
|  | Delay vs normal | 1.04 | 0.74 - 1.41 | 1.01 | 0.58 - 1.62 | 1.82 | 1.09 - 2.85 | 2.48 | 0.88 - 5.41 | 1.65 | 0.90 - 2.77 |
|  | Catch-up vs delay | 1.01 | 0.89 - 1.14 | 1.01 | 0.83 - 1.21 | 0.94 | 0.74 - 1.17 | 0.95 | 0.61 - 1.40 | 0.93 | 0.70 - 1.21 |
| **Personal/social** |  |  |  |  |  |  |  |  |  |  |  |
|  | Catch-up vs normal | 1.07 | 0.95 - 1.21 | 1.10 | 0.91 - 1.33 | 1.17 | 0.92 - 1.46 | 1.56 | 0.99 - 2.31 | 1.06 | 0.80 - 1.38 |
|  | Delay vs normal | 1.09 | 0.87 - 1.35 | 1.02 | 0.71 - 1.43 | 1.32 | 0.87 - 1.90 | 1.50 | 0.60 - 3.07 | 1.27 | 0.80 - 1.91 |
|  | Catch-up vs delay | 0.99 | 0.87 - 1.12 | 1.01 | 0.83 - 1.22 | 0.97 | 0.76 - 1.21 | 1.02 | 0.65 - 1.51 | 0.95 | 0.71 - 1.23 |

Reference group: HDP-unaffected. Adjusted for maternal age at pregnancy, family annual income, maternal educational attainment, maternal pre-pregnancy body mass index, parity, gestational diabetes mellitus prevalence, maternal tobacco use during pregnancy, maternal alcohol use during pregnancy, maternal folic acid intake during pregnancy, and the child’s sex. HDP, hypertensive disorders of pregnancy; PE, preeclampsia; GH, gestational hypertension; EO, early onset; LO, late onset; RR, risk ratio; CI, confidence interval.

**Supplementary Table 6-2. Risk differences in child developmental patterns across HDP subtypes after excluding congenital abnormalities**

| **Domain** | **Pattern comparison** | **HDP** | | **GH** | | **PE** | | **PEEO** | | **PELO** | |
| --- | --- | --- | --- | --- | --- | --- | --- | --- | --- | --- | --- |
|  |  | **Adjusted RR** | **95% CI** | **Adjusted RR** | **95% CI** | **Adjusted RR** | **95% CI** | **Adjusted RR** | **95% CI** | **Adjusted RR** | **95% CI** |
| **Communication** |  |  |  |  |  |  |  |  |  |  |  |
|  | Catch-up vs normal | 1.06 | 0.94 - 1.19 | 1.02 | 0.85 - 1.21 | 1.16 | 0.93 - 1.42 | 1.55 | 1.04 - 2.22 | 1.04 | 0.80 - 1.33 |
|  | Delay vs normal | 1.19 | 0.99 - 1.42 | 1.04 | 0.77 - 1.38 | 1.44 | 1.03 - 1.95 | 2.02 | 1.10 - 3.35 | 1.27 | 0.84 - 1.82 |
|  | Catch-up vs delay | 0.96 | 0.85 - 1.07 | 0.99 | 0.82 - 1.17 | 0.93 | 0.74 - 1.14 | 0.93 | 0.62 - 1.33 | 0.93 | 0.71 - 1.19 |
| **Gross motor** |  |  |  |  |  |  |  |  |  |  |  |
|  | Catch-up vs normal | 1.06 | 0.93 - 1.21 | 0.98 | 0.79 - 1.20 | 1.22 | 0.95 - 1.53 | 1.66 | 1.06 - 2.46 | 1.09 | 0.81 - 1.43 |
|  | Delay vs normal | 1.28 | 1.07 - 1.52 | 1.22 | 0.92 - 1.58 | 1.61 | 1.17 - 2.14 | 2.01 | 1.10 - 3.34 | 1.49 | 1.02 - 2.08 |
|  | Catch-up vs delay | 0.93 | 0.82 - 1.07 | 0.92 | 0.74 - 1.13 | 0.91 | 0.71 - 1.15 | 0.93 | 0.59 - 1.38 | 0.90 | 0.67 - 1.18 |
| **Fine motor** |  |  |  |  |  |  |  |  |  |  |  |
|  | Catch-up vs normal | 1.11 | 1.00 - 1.24 | 1.15 | 0.98 - 1.34 | 1.12 | 0.91 - 1.37 | 1.25 | 0.83 - 1.80 | 1.09 | 0.85 - 1.36 |
|  | Delay vs normal | 1.19 | 0.86 - 1.62 | 0.84 | 0.45 - 1.43 | 1.80 | 1.04 - 2.89 | 2.72 | 1.07 - 5.63 | 1.50 | 0.74 - 2.67 |
|  | Catch-up vs delay | 1.00 | 0.90 - 1.11 | 1.03 | 0.88 - 1.20 | 0.96 | 0.78 - 1.16 | 0.91 | 0.60 - 1.31 | 0.97 | 0.76 - 1.22 |
| **Problem solving** |  |  |  |  |  |  |  |  |  |  |  |
|  | Catch-up vs normal | 1.13 | 1.00 - 1.26 | 1.06 | 0.88 - 1.27 | 1.18 | 0.94 - 1.46 | 1.66 | 1.12 - 2.36 | 1.03 | 0.78 - 1.33 |
|  | Delay vs normal | 1.11 | 0.81 - 1.48 | 0.92 | 0.54 - 1.48 | 1.92 | 1.18 - 2.95 | 2.96 | 1.26 - 5.83 | 1.63 | 0.88 - 2.72 |
|  | Catch-up vs delay | 1.00 | 0.89 - 1.13 | 1.01 | 0.84 - 1.21 | 0.93 | 0.74 - 1.15 | 0.92 | 0.62 - 1.31 | 0.93 | 0.70 - 1.20 |
| **Personal/social** |  |  |  |  |  |  |  |  |  |  |  |
|  | Catch-up vs normal | 1.07 | 0.95 - 1.21 | 1.06 | 0.88 - 1.27 | 1.17 | 0.93 - 1.45 | 1.57 | 1.04 - 2.24 | 1.04 | 0.79 - 1.35 |
|  | Delay vs normal | 1.16 | 0.94 - 1.42 | 1.07 | 0.76 - 1.46 | 1.41 | 0.95 - 1.99 | 1.50 | 0.64 - 2.93 | 1.38 | 0.88 - 2.04 |
|  | Catch-up vs delay | 0.98 | 0.87 - 1.10 | 0.99 | 0.82 - 1.18 | 0.96 | 0.77 - 1.19 | 1.04 | 0.69 - 1.49 | 0.93 | 0.70 - 1.20 |

Reference group: HDP-unaffected. Adjusted for maternal age at pregnancy, family annual income, maternal educational attainment, maternal pre-pregnancy body mass index, parity, gestational diabetes mellitus prevalence, maternal tobacco use during pregnancy, maternal alcohol use during pregnancy, maternal folic acid intake during pregnancy, and the child’s sex. HDP, hypertensive disorders of pregnancy; PE, preeclampsia; GH, gestational hypertension; EO, early onset; LO, late onset; RR, risk ratio; CI, confidence interval.
